# Environment-Wide Association Study of Chemical Biomarkers and Health Outcomes in NHANES 2017–2018: Discovery, Validation, and Dose–Response Analysis

**DOI:** 10.64898/2026.02.07.26345792

**Authors:** Hayden Farquhar

## Abstract

**Background:** Environment-wide association studies (ExWAS) offer a systematic approach to identifying chemical biomarker–health outcome associations, yet few have applied rigorous multi-stage validation.

**Methods:** We screened 92 chemical biomarkers against 48 health outcomes in NHANES 2017–2018 (2,796 tests across four screening rounds; not all chemicals were crossed with all outcomes). Associations passing an initial FDR screen were subjected to cross-cycle validation in NHANES 2015–2016—the primary inferential safeguard given the adaptive screening design—followed by dose–response analysis and multiple sensitivity specifications. Survey-weighted regression models adjusted for age, sex, race/ethnicity, poverty–income ratio, BMI, and smoking.

**Results:** Of 26 associations passing FDR correction, 21 were testable in cross-cycle validation; of these, 15 (71%) replicated with concordant direction and p < 0.05 in a temporally independent NHANES 2015–2016 sample. Of these 15, 14 remained robust after analyte-specific sensitivity checks; urinary creatinine adjustment identified one association (iodine–BMI) as a dilution artifact. Two novel findings emerged: dimethylarsonic acid with uric acid (β = 0.20 mg/dL per log-unit DMA, 95% CI: 0.15–0.26) and urinary perchlorate with BUN (β = 1.21 mg/dL per log-unit perchlorate, 95% CI: 0.97–1.45); a third high-novelty association (methylmercury–waist circumference) is likely explained by fish consumption patterns.

**Conclusions:** Multi-stage ExWAS with cross-cycle validation identified 14 robust chemical–health associations. Two novel findings—DMA–uric acid and perchlorate–BUN—survived all sensitivity checks and warrant prospective investigation.

## 1 Introduction

Environmental chemical exposures are ubiquitous in the general population and have been implicated in a broad spectrum of adverse health outcomes, from metabolic and cardiovascular disease to neurodevelopmental and reproductive disorders. However, most environmental epidemiology studies adopt a hypothesis-driven approach, testing one or a small number of pre-specified chemical–outcome pairs. This limits the ability to detect unexpected associations and introduces publication bias favoring positive results for well-studied chemicals.

The environment-wide association study (ExWAS) paradigm, first proposed by Patel et al. [1], offers a complementary, hypothesis-free approach that systematically screens many chemical exposures against health outcomes in a single analytical framework. By analogy to genome-wide association studies (GWAS), ExWAS leverages large biomonitoring datasets to identify associations that merit further investigation while controlling for multiple comparisons.

The National Health and Nutrition Examination Survey (NHANES) is well suited for ExWAS. It provides nationally representative biomonitoring data on hundreds of chemical exposures—heavy metals, phthalates, perfluoroalkyl substances, volatile organic compounds, pesticides, and others—alongside clinical laboratory panels and examination data, all within a complex survey design that permits population-level inference.

Despite this resource, most published ExWAS analyses have been limited in scope, often restricted to a single outcome domain or chemical class, and few have applied multi-stage validation. The critical gap is replication: associations identified in a discovery phase are rarely tested in independent data, leaving open whether they represent true biological relationships or statistical artifacts of multiple testing.

We conducted a comprehensive ExWAS using NHANES 2017–2018, screening 2,796 exposure–outcome combinations across 92 chemical biomarkers and 48 health outcomes. Findings that survived false discovery rate correction were subjected to a four-stage validation pipeline: (1) cross-cycle replication in NHANES 2015–2016, (2) quartile-based dose–response analysis, (3) nine sensitivity specifications testing robustness to analytic decisions, and (4) structured literature review for novelty assessment. This multi-stage approach minimizes the risk of false positives inherent to high-dimensional screening while identifying genuinely novel associations for targeted follow-up.

## 2 Methods

### 2.1 Study Population

We used data from the National Health and Nutrition Examination Survey (NHANES) 2017–2018 cycle, a nationally representative, cross-sectional survey of the non-institutionalized civilian US population conducted by the National Center for Health Statistics (NCHS). NHANES employs a complex, multistage probability sampling design with oversampling of certain demographic groups [2,3]. All participants provided written informed consent, and the protocol was approved by the NCHS Research Ethics Review Board. Our analysis was restricted to adults aged 18 years and older with valid survey weights and non-missing covariate data. Of the NHANES 2017–2018 participants examined at the mobile examination center, 5,265 adults met these inclusion criteria and formed the core analytic sample for blood-based biomarker analyses (WTMEC2YR weights). Urinary subsample A analyses were restricted to approximately 1,580 adults, and surplus serum analyses to approximately 1,370 adults, reflecting the smaller subsample designs of those NHANES components. This study was not pre-registered; the analysis is exploratory, consistent with the ExWAS paradigm, but employs a multi-stage validation architecture (FDR correction, cross-cycle replication, dose–response analysis, sensitivity specifications) designed to distinguish robust signals from statistical artifacts. The validation protocol—including cross-cycle replication in 2015–2016, dose–response analysis, and the nine sensitivity specifications—was specified before examining any 2015–2016 data.

The analytic sample comprised approximately 4,800–5,300 participants depending on outcome and exposure subsample, representing an estimated 218 million non-institutionalized US adults. The mean age was 48.2 years (SD = 17.1), 51.7% were female, 63.1% were non-Hispanic White, 10.8% were non-Hispanic Black, and 26.1% were other race/ethnicity groups. Mean BMI was 29.8 kg/m^2^ (SD = 7.1), and 23.4% were current smokers.

**Table 1.** Demographic and clinical characteristics of the study population.

| Characteristic | Unweighted n | Weighted estimate |
| --- | --- | --- |
| Sample size | 5,265 | 218,119,343 (population) |
| Age, mean (SD) | – | 48.2 (17.1) years |
| Female, % | 2,713 (51.5%) | 51.7% |
| Race/ethnicity |  |  |
| – Non-Hispanic White | 1,812 (34.4%) | 63.1% |
| – Non-Hispanic Black | 1,259 (23.9%) | 10.8% |
| – Other | 2,194 (41.7%) | 26.1% |
| Poverty-income ratio, mean (SD) | – | 3.00 (1.58) |
| BMI, mean (SD) | – | 29.8 (7.1) kg/m<sup>2</sup> |
| Current smoker, % | 1,231 (23.4%) | 23.4% |
| Waist circumference, mean (SD) | – | 101.0 (17.3) cm |

Note: Unweighted percentages reflect NHANES oversampling design; weighted estimates represent the US adult population.

### 2.2 Chemical Exposure Biomarkers

We measured 92 chemical biomarkers spanning nine chemical classes: heavy metals (blood lead, cadmium, mercury, selenium, manganese, cobalt), per- and polyfluoroalkyl substances (PFAS; PFNA, PFHxS, PFDeA, PFUA), phthalate metabolites (MEHHP, MEOHP, MECPP, and others), polycyclic aromatic hydrocarbon (PAH) metabolites, volatile organic compound (VOC) metabolites, urinary metals and elements (iodine, perchlorate, cesium, thallium, arsenic species), pesticides (glyphosate, oxychlordane), and other environmental chemicals.

All chemical concentrations were measured in NHANES laboratories following standardized protocols. Values below the limit of detection (LOD) were replaced with LOD/^√^2, a standard approach for left-censored environmental data [4,5]. Chemicals with greater than 70% of values at or below the LOD were excluded from analysis due to insufficient variability for regression modeling. We used a 70% threshold rather than the more restrictive 40–50% cutoffs sometimes applied in single-chemical studies, in order to maximize the breadth of the chemical screen while excluding only chemicals whose exposure distributions would be dominated by the LOD/^√^2 pile-up. None of the 15 validated findings involved chemicals near this exclusion threshold, as the implicated biomarkers (blood lead, selenium, manganese, mercury, urinary DMA, perchlorate, iodine) all had detection frequencies exceeding 95%. Of the 92 chemicals screened, 12 had detection frequencies between 40–70%; excluding these would reduce the test count from 2,796 to 2,220 but does not change the FDR-significant findings (Table S9). All exposure biomarkers were natural-log-transformed prior to regression to reduce right skewness and improve model fit.

### 2.3 Health Outcomes

We examined 48 health outcomes across seven clinical domains: metabolic markers (total cholesterol, HDL, triglycerides, HbA1c, fasting glucose, uric acid), liver enzymes (ALT, AST, GGT, alkaline phosphatase [ALP], total bilirubin), kidney function (eGFR, blood urea nitrogen), hematological indices (RBC count, hemoglobin, WBC, platelets), anthropometric measures (BMI, waist circumference), cardiovascular (systolic and diastolic blood pressure), and mental health (PHQ-9 depression score). These seven domains comprised 24 outcomes used in the broad and expanded screening rounds. The full complement of 48 reflects the addition of 10 thyroid function markers (free T3, free T4, total T3, total T4, TSH, thyroglobulin, and thyroid antibodies, tested in the PFAS–thyroid screen) and 14 blood chemistry markers including HDL cholesterol, chloride, sodium, potassium, phosphorus, total protein, and bicarbonate (tested in the novelty screen). Estimated glomerular filtration rate (eGFR) was calculated using the race-free 2021 CKD-EPI creatinine-only equation [6], as cystatin C was not measured in all NHANES 2017–2018 participants.

### 2.4 Statistical Analysis

#### 2.4.1 Discovery phase

Survey-weighted linear regression models were fit for each exposure–outcome pair using svyglm() from the R survey package [7,8], accounting for the complex survey design (clustering by primary sampling unit, stratification, and appropriate survey weights). The covariate model included:

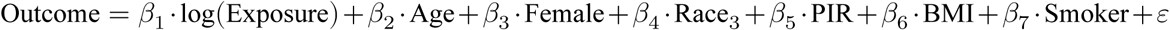

where Race_3_ was a three-level variable (Non-Hispanic White, Non-Hispanic Black, Other) collapsed from the six-level NHANES RIDRETH3 variable to ensure model stability in subsample analyses; a six-level sensitivity analysis confirmed minimal impact on effect estimates (Table S7). PIR is the poverty–income ratio, BMI is body mass index (omitted when BMI or waist circumference was the outcome to avoid collider bias; effect sizes for anthropometric outcomes are therefore not directly comparable to those for other outcomes), and Smoker is a binary indicator for current smoking. The covariate set was selected a priori to capture the primary confounding pathways between chemical exposures and health outcomes: age (cumulative exposure and physiological changes), sex (metabolic differences and exposure patterns), race/ethnicity (socioeconomic and dietary factors), PIR (residential and occupational exposure gradients), BMI (metabolic confounding), and smoking (a major chemical exposure source and independent risk factor). Alcohol consumption was not included in the primary model due to extensive missingness from NHANES ALQ_J skip patterns (only 3,143 of 5,265 blood biomarker participants and 1,035 of approximately 1,580 urinary subsample participants had complete alcohol data); alcohol adjustment was instead conducted as a sensitivity analysis (see below). This parsimonious specification preserves degrees of freedom within the NHANES complex survey design while addressing the most important confounders. A directed acyclic graph (DAG) illustrating the causal assumptions underlying this covariate selection is provided in Figure S18; the DAG was constructed a priori based on established confounding pathways in environmental epidemiology, not fitted to the data. Survey weights were selected based on exposure source: WTMEC2YR for blood-based biomarkers (LBX prefix), WTSA2YR for urinary subsample A biomarkers (URX prefix), and WTSSBJ2Y for surplus serum samples (SS prefix).

#### 2.4.2 Multiple testing correction

We applied the Benjamini–Hochberg FDR procedure [9] globally across all 2,796 tests, pooling p-values from analyses using different survey weights (WTMEC2YR for blood biomarkers, WTSA2YR for urinary subsample, WTSSBJ2Y for surplus serum) into a single ranked list. This pooling is methodologically defensible because FDR control depends only on the validity of individual p-values under the null, not on comparable sample sizes across tests; however, it does mean that urinary and surplus serum findings face implicitly higher thresholds due to lower power. Associations with FDR < 0.05 were considered statistically significant and advanced to the validation stage.

The four screening rounds were conducted sequentially rather than simultaneously: the PFAS–thyroid and broad screens were hypothesis-driven, while the expanded and novelty screens broadened the chemical– outcome space after initial null results. This adaptive design inflates the effective number of comparisons beyond what the Benjamini–Hochberg procedure formally controls; **FDR-corrected p-values should therefore be interpreted as descriptive summaries of relative signal strength rather than inferential error rates.** Within-round FDR correction is reported in Table S6 for comparison. **Cross-cycle validation in the independent 2015–2016 NHANES cycle—not FDR correction—serves as the primary inferential safeguard against false positives.**

#### 2.4.3 Cross-cycle validation

FDR-significant associations were replicated in a temporally independent NHANES 2015–2016 cycle. The identical covariate model and survey design specifications were applied. An association was considered validated if the effect estimate was in the same direction as the discovery analysis and the replication p-value was < 0.05. Associations involving chemicals not measured in the 2015–2016 cycle (e.g., glyphosate surplus serum) or where models failed to converge were classified as untestable. We note that consecutive NHANES cycles share methodological infrastructure (laboratory protocols, questionnaire instruments, sampling frame) and draw from the same underlying US population, so cross-cycle replication tests temporal stability of associations rather than true external generalizability to different populations or measurement systems. This is a limitation common to all NHANES-based ExWAS studies.

#### 2.4.4 Dose–response analysis

For validated associations, we categorized exposure into quartiles and estimated the mean outcome at each quartile level using survey-weighted regression, adjusting for the same covariates. Trend significance was assessed using a linear contrast across quartile midpoints. Monotonicity was evaluated by inspecting whether effect estimates increased (or decreased) consistently across quartiles.

#### 2.4.5 Sensitivity analyses

A comprehensive battery of sensitivity analyses was applied to each validated finding, comprising 9 primary specifications and multiple additional robustness checks. The nine primary specifications were: (1) female-only subgroup, (2) male-only subgroup, (3) age < 50 years, (4) age ≥ 50 years, (5) exclusion of outliers beyond the 99th percentile, (6) adjustment for education (additional covariate), (7) substitution of serum cotinine for binary smoking, (8) restriction to adults aged 20 years and older, and (9) the primary model (as reference). A finding was considered robust if the direction of association was concordant and the p-value was < 0.05 in at least 7 of 9 primary specifications (permitting up to two failures to accommodate expected power loss in sex- or age-stratified subgroups, which halve the effective sample size).

Four additional sensitivity specifications targeted specific confounding concerns: (1) adjustment for fish consumption (number of fish/shellfish meals in the past 30 days, NHANES variable DBD895) for all findings, (2) adjustment for moderate-to-vigorous recreational physical activity (derived from PAQ_J: participants meeting World Health Organization [WHO] guidelines of ≥ 150 minutes/week moderate-equivalent activity), (3) adjustment for alcohol consumption (derived from ALQ_J frequency and quantity variables), and (4) adjustment for urinary creatinine (log-transformed, from ALB_CR_J) for urinary biomarker associations to address dilution variation [10]. For the DMA–uric acid and perchlorate–BUN findings specifically, additional adjustment for total protein intake (grams/day from 24-hour dietary recall, DR1TPROT) was conducted to address diet as a potential confounder for these metabolic outcomes (Table S8). A sensitivity analysis using the full six-level NHANES RIDRETH3 race/ethnicity classification (Mexican American, Other Hispanic, Non-Hispanic White, Non-Hispanic Black, Non-Hispanic Asian, Other/Multiracial) was conducted for blood biomarker findings where sample sizes supported the finer categorization (Table S7). Additional model specifications included quadratic age adjustment (Table S13) and 24-hour dietary recall fish consumption for mercury findings (Table S12).

#### 2.4.6 Post-hoc power analysis

Post-hoc power analysis was conducted using Cohen’s *f* ^2^ framework with 80% power, *α* adjusted for multiple comparisons (Bonferroni and FDR thresholds), and a design effect of 2 to account for NHANES complex sampling. Effective sample sizes were halved from raw counts to approximate the variance inflation from clustering and stratification.

#### 2.4.7 Novelty assessment

A structured PubMed literature search was conducted for each validated finding using chemical and outcome keywords, supplemented with Medical Subject Headings (MeSH)-based searches for HIGH-novelty findings (Table S14). Embase, Scopus, and Web of Science were not searched; for the two genuinely novel findings (DMA–uric acid, perchlorate–BUN), this represents a limitation, though major environmental epidemiology studies are typically indexed in PubMed. Findings were classified as HIGH novelty (0–2 relevant publications, no direct prior evidence), MODERATE (1–8 publications, partially consistent), LOW-MODERATE (limited literature, consistent direction), or LOW (≥ 20 publications, well-established).

#### 2.4.8 Software

All analyses were performed in R (version 4.3) using the survey package for complex survey regression, nhanesA for programmatic NHANES data access, tidyverse for data manipulation, and broom for model output extraction. Figures were generated with ggplot2, ggrepel, forestplot, and pheatmap. Analysis scripts and version details are available at https://github.com/hayden-farquhar/NHANES-EXWAS.

## 3 Results

### 3.1 Discovery Phase

We conducted 2,796 survey-weighted regression tests across four screening rounds, summarized below:

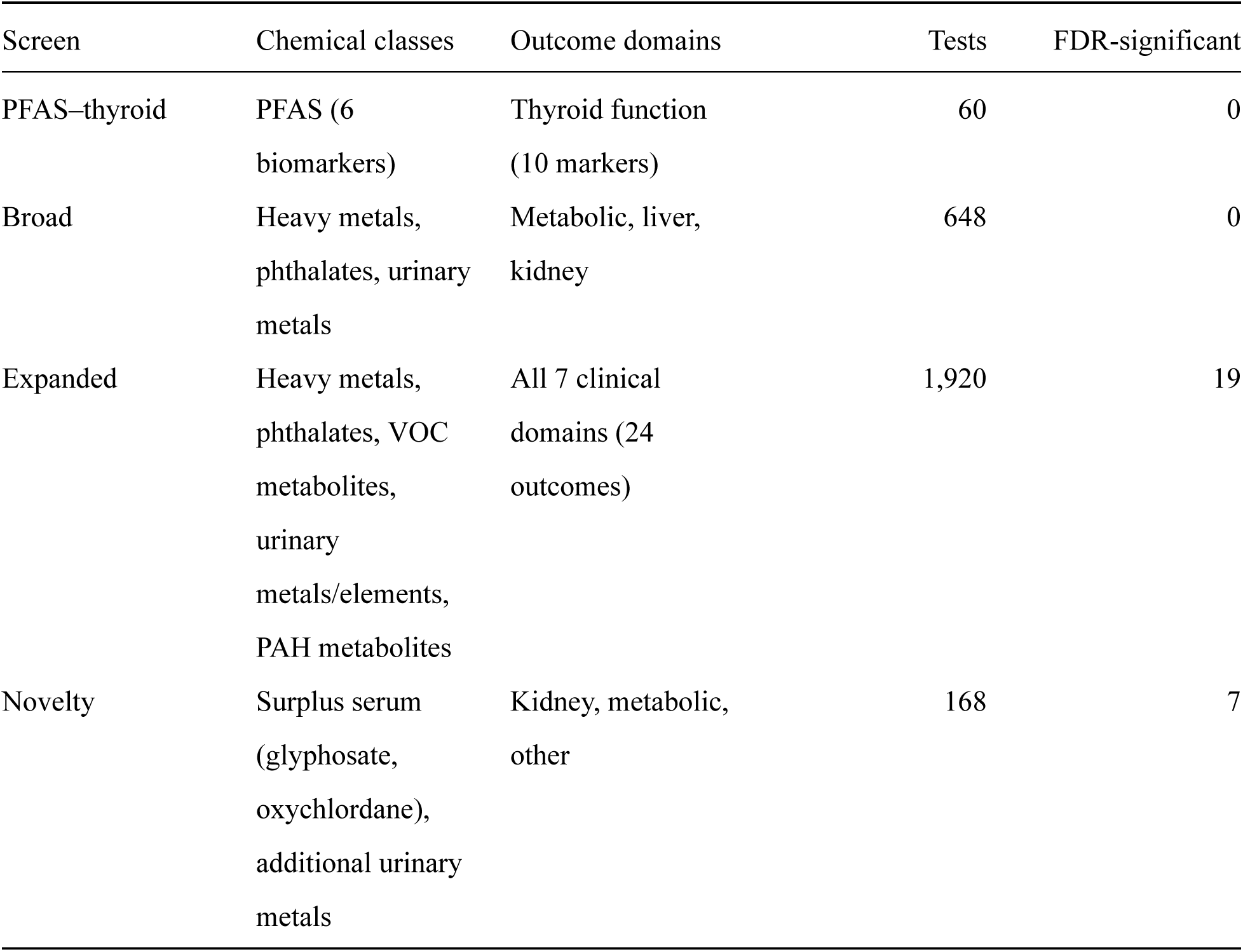

The PFAS–thyroid and broad screens yielded no FDR-significant associations. All 26 significant findings (after global Benjamini–Hochberg correction) emerged from the expanded screen—which systematically crossed 80 chemicals with 24 outcomes—and the novelty screen, which targeted surplus serum analytes and chemical–outcome pairs that showed suggestive signals (p < 0.01 but FDR > 0.05) in the 2017–2018 discovery phase. This sequential design—where the novelty screen was informed by preliminary examination of the discovery data—could inflate type I error if validation relied solely on FDR correction. The critical safeguard is that all findings were subsequently tested in the temporally independent 2015–2016 NHANES cycle, which was not examined until after the discovery-phase screens were finalized. Cross-cycle validation, not FDR correction, provides the primary protection against false positives arising from this data-adaptive design. The 92 unique chemicals comprised the 80 tested in the expanded screen, 6 PFAS biomarkers tested exclusively against thyroid markers, and 6 surplus serum and additional urinary analytes tested in the novelty screen. These 26 associations involved 17 unique chemical biomarkers and 13 unique health outcomes (Table S1; Figure 1). One association (blood manganese with RBC count) also met the Bonferroni threshold.

**Figure 1:**
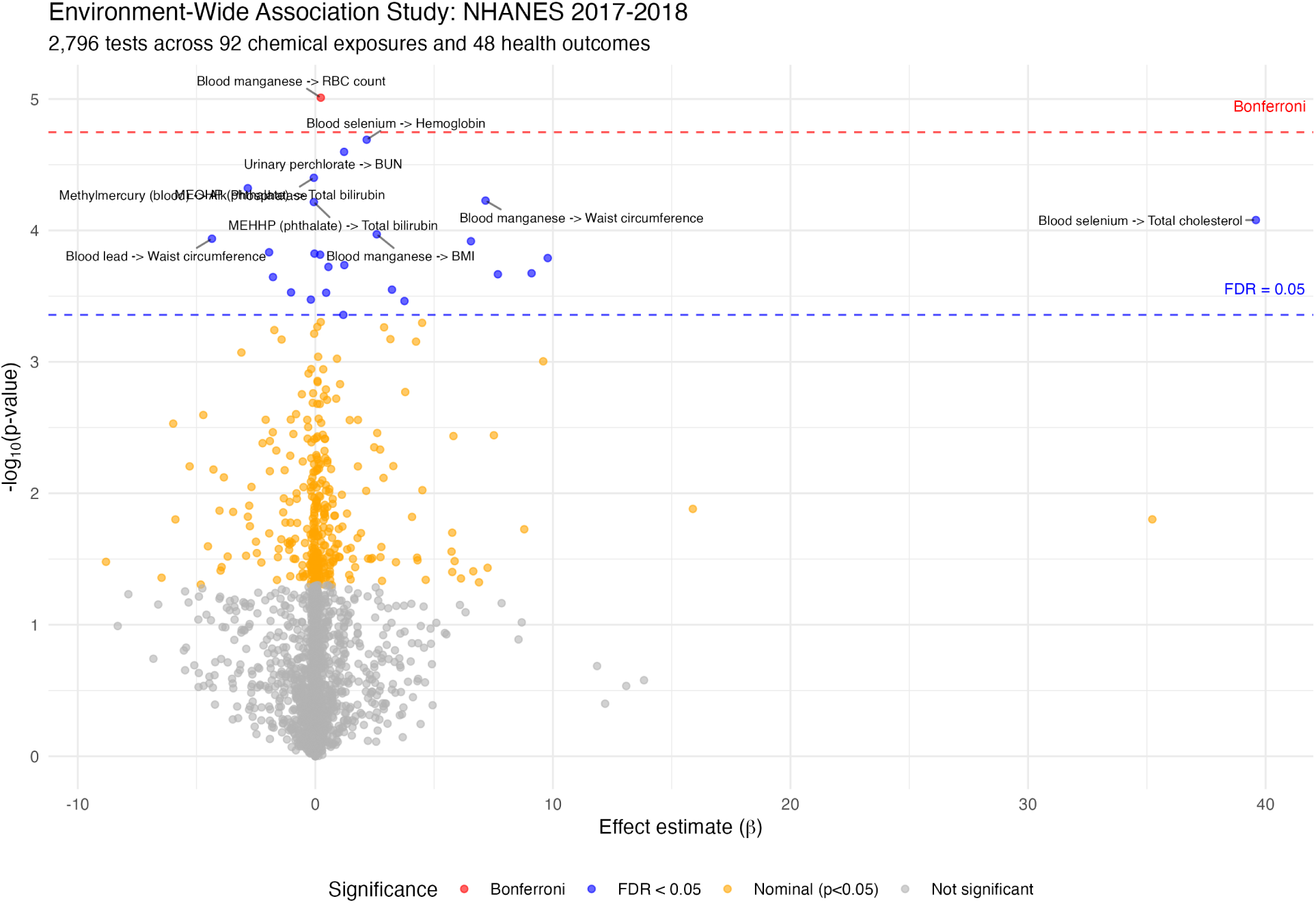
Volcano plot of all 2,796 exposure–outcome associations. The x-axis shows unstandardized β coefficients, which are not directly comparable across outcomes measured in different units; the plot is intended to show the distribution of effect directions and significance levels rather than relative effect magnitudes. Points above the dashed line exceed FDR < 0.05; labeled points identify the strongest signals by p-value. Standardized effect size (t/√n) and partial R² versions of this plot are provided in Figures S16–S17 for direct comparison of effect magnitudes across outcomes.

**Figure 2:**
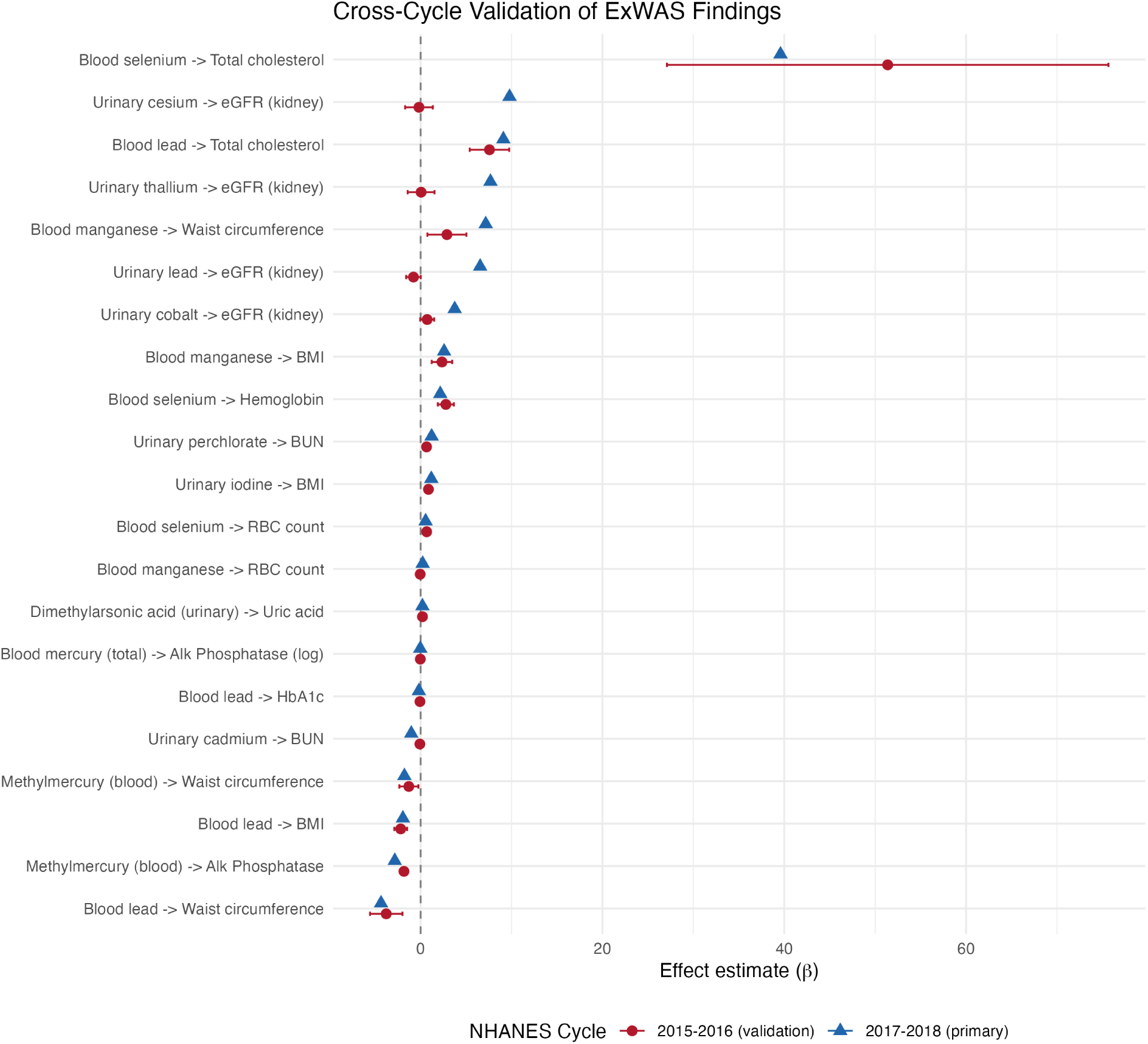
Forest plot of 21 testable FDR-significant associations showing discovery (2017–2018) and validation (2015–2016) estimates with 95% confidence intervals. Associations are ordered by validation p-value; the 15 that met validation criteria (concordant direction, p < 0.05) appear at the top.

The strongest association was blood manganese (LBXBMN) positively associated with RBC count (LBXR- BCSI; *β* = 0.23 × 10^12^ cells/L per log-unit increase, 95% CI: 0.19–0.27, p = 9.8 × 10^-6^, FDR = 0.024, n = 4,873), the only result that also cleared the Bonferroni threshold. Other prominent signals included blood selenium (LBXBSE) with hemoglobin (*β* = 2.16 g/dL, 95% CI: 1.74–2.58, FDR = 0.024), urinary perchlorate (URXUP8) with BUN (*β* = 1.21 mg/dL, 95% CI: 0.97–1.45, FDR = 0.024), and methylmercury (LBXBGM) with ALP (*β* = −2.84 U/L, 95% CI: −3.47 to −2.21, FDR = 0.024).

Heavy metals dominated the significant findings (13 of 26 associations), followed by urinary metals/elements and phthalates. The most frequently implicated outcome domains were anthropometric, metabolic, and kidney function markers. Full results for all 26 FDR-significant associations are provided in Table S1. Note that many FDR-corrected q-values cluster at similar values (e.g., 0.024, 0.030); this is expected behavior of the Benjamini–Hochberg procedure, which assigns the same adjusted p-value to all tests sharing the same rank-order position relative to the FDR threshold. Because the volcano plot (Figure 1) displays unstandardized *β* coefficients on the x-axis, effect sizes are not directly comparable across outcomes measured in different units; Figures S16–S17 present standardized versions (t/√n and partial R²) that enable cross-outcome comparison.

### 3.2 Cross-Cycle Validation

The 26 FDR-significant associations from the discovery phase were advanced to cross-cycle validation in NHANES 2015–2016.

Of the 26 FDR-significant associations, 5 could not be tested in the 2015–2016 cycle: 2 involved phthalate metabolites where models failed to converge, 2 involved glyphosate (not measured in the 2015–2016 surplus serum panel), and 1 involved oxychlordane (not available). Of the remaining 21 testable associations, 15 (71%) were validated with concordant direction and p < 0.05 in the 2015–2016 cycle (Table 2).

**Table 2.**
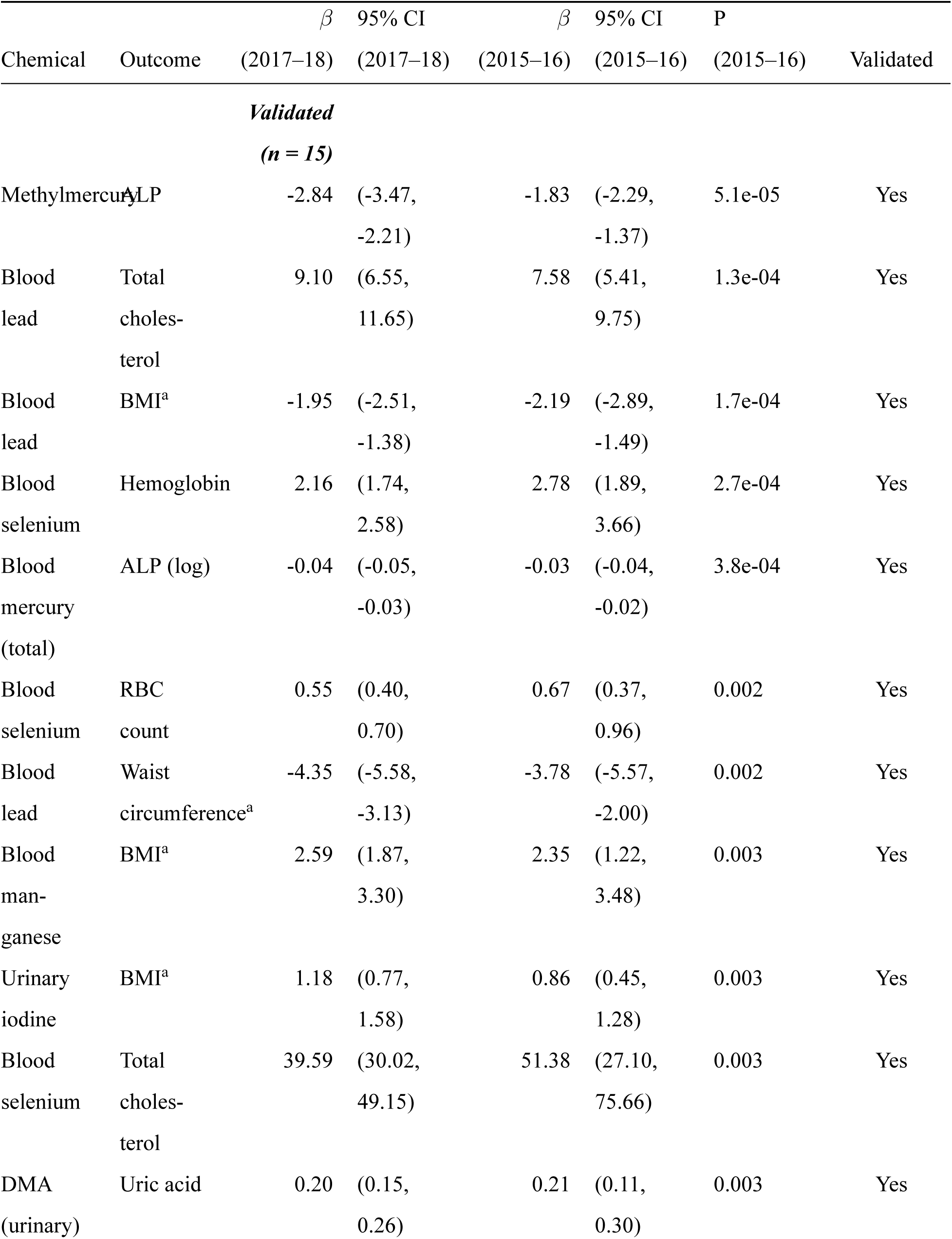
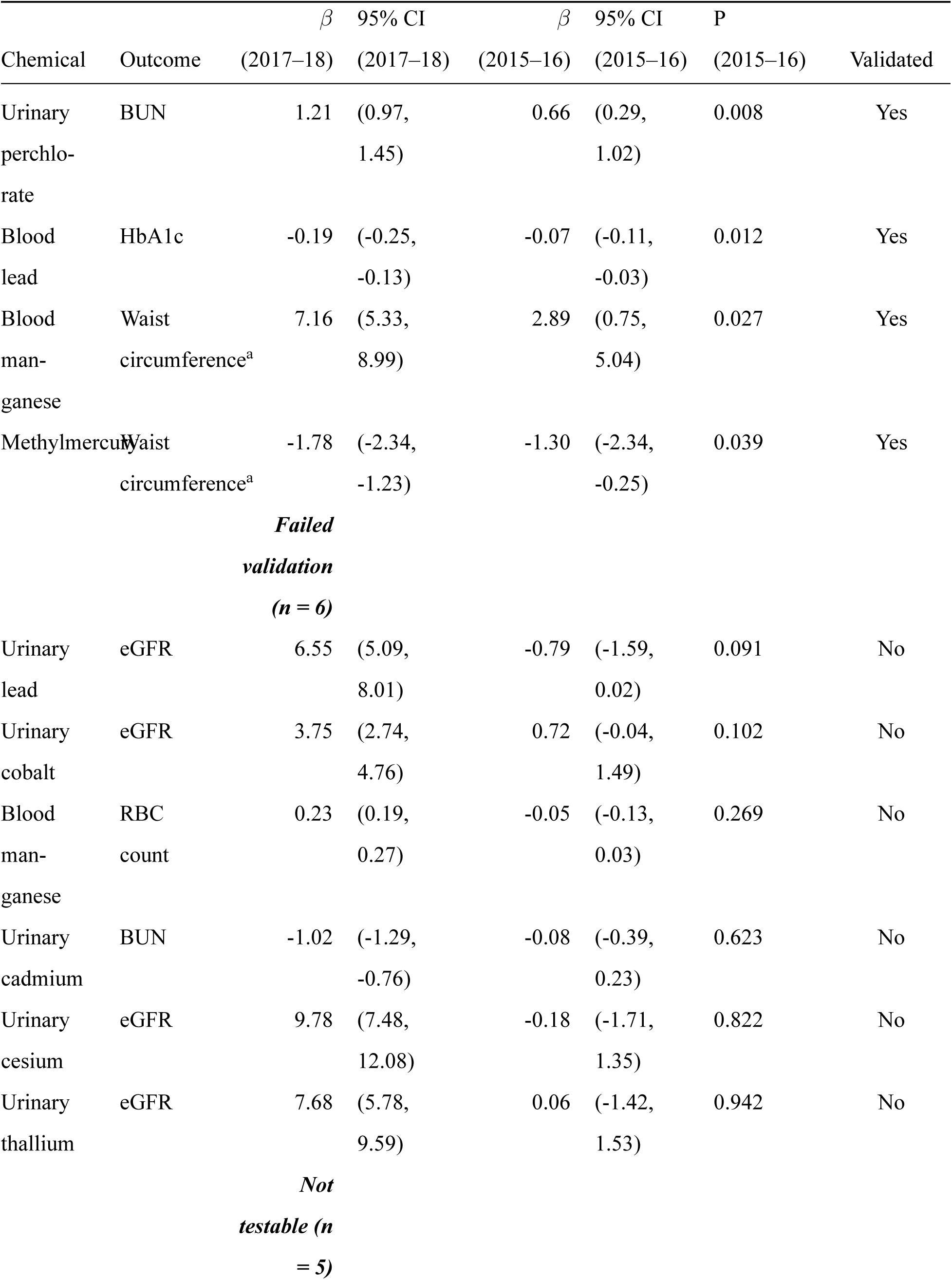
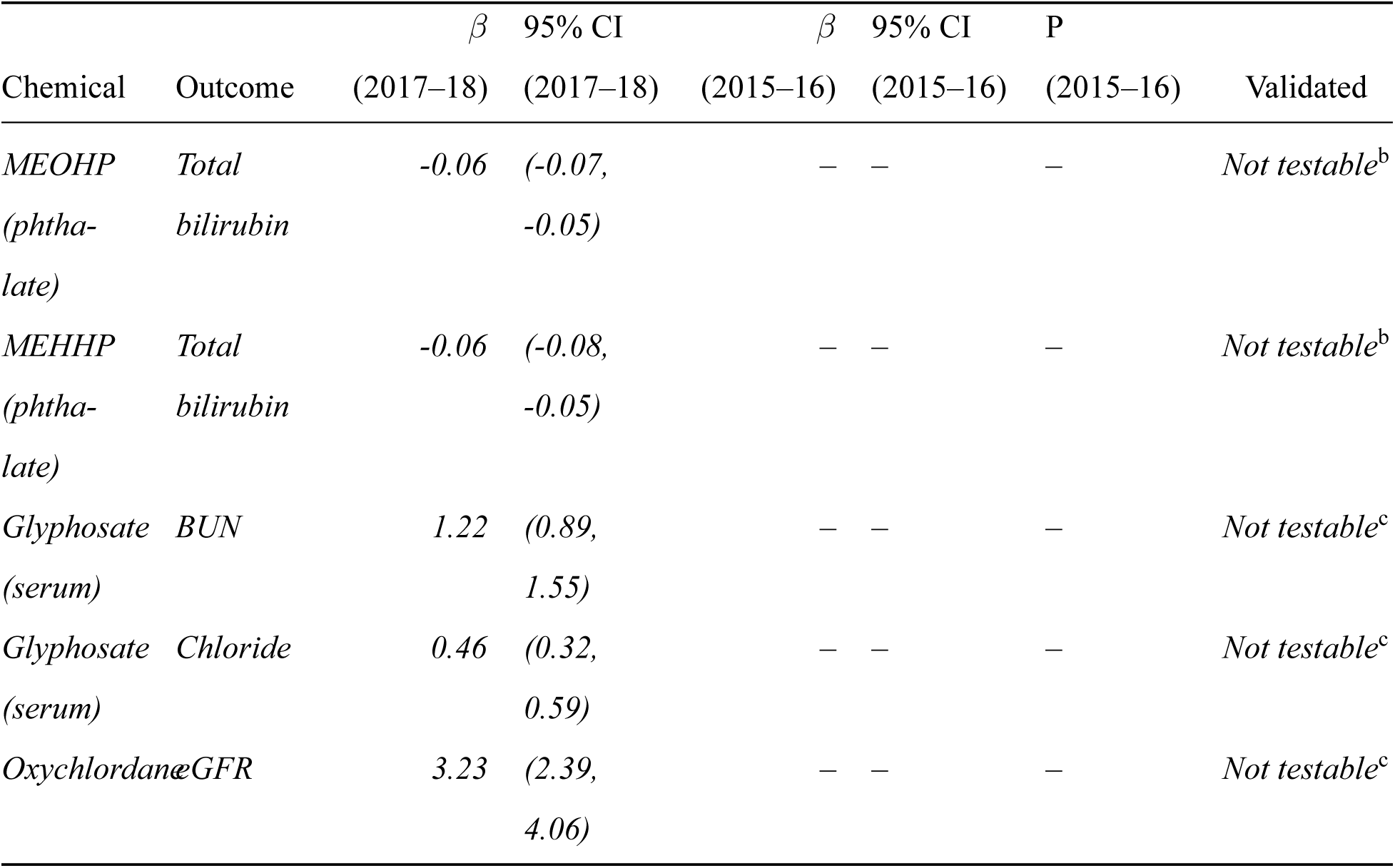
Cross-cycle validation results for all 26 FDR-significant associations. ^a^Models for BMI and waist circumference outcomes omit BMI from the covariate set to avoid collider bias. ^b^Not testable due to model convergence failure (phthalate–bilirubin models in the 2015–2016 cycle). ^c^Not testable due to chemical not measured in 2015–2016 (glyphosate surplus serum panel; oxychlordane).

Validated associations included:

- **Blood lead** with BMI (*β*_2017_ = −1.95, *β*_2015_ = −2.19, p = 1.7 × 10^-4^), waist circumference, total cholesterol, and HbA1c
- **Blood selenium** with hemoglobin (*β*_2017_ = 2.16, *β*_2015_ = 2.78, p = 2.7 × 10^-4^), RBC count, and total cholesterol
- **Methylmercury** with ALP (*β*_2017_ = −2.84, *β*_2015_ = −1.83, p = 5.1 × 10^-5^) and waist circumference
- **Blood manganese** with BMI and waist circumference
- **Dimethylarsonic acid** with uric acid (*β*_2017_ = 0.20, *β*_2015_ = 0.21, p = 0.003)
- **Urinary perchlorate** with BUN (*β*_2017_ = 1.21, *β*_2015_ = 0.66, p = 0.008)
- **Urinary iodine** with BMI (*β*_2017_ = 1.18, *β*_2015_ = 0.86, p = 0.003)
- **Blood mercury (total)** with ALP (log-transformed)

Six associations failed validation, including blood manganese with RBC count (the Bonferroni-significant finding in 2017–2018 that showed a reversed direction in 2015–2016), and four urinary metal–eGFR associations (cesium, thallium, lead, cobalt), suggesting these kidney findings may be cycle-specific or susceptible to unmeasured confounding. An additional five associations could not be tested: two phthalate–bilirubin models failed to converge in the validation cycle, two glyphosate associations could not be tested because glyphosate was not measured in the 2015–2016 surplus serum panel, and one oxychlordane–eGFR association lacked the requisite exposure data. The 71% validation rate among testable associations compares favorably with prior ExWAS replication studies; for example, Patel et al. [11] reported that approximately half of environment–mortality associations replicated across NHANES cycles.

### 3.3 Dose–Response Relationships

All 15 validated associations showed statistically significant dose–response trends in quartile analyses (p_trend_ range: 7.7 × 10^-5^ to 4.3 × 10^-3^). Of these, 14 exhibited monotonic dose–response gradients, with effect estimates increasing (or decreasing) consistently from the first to fourth quartile (Figure 3).

**Figure 3:**
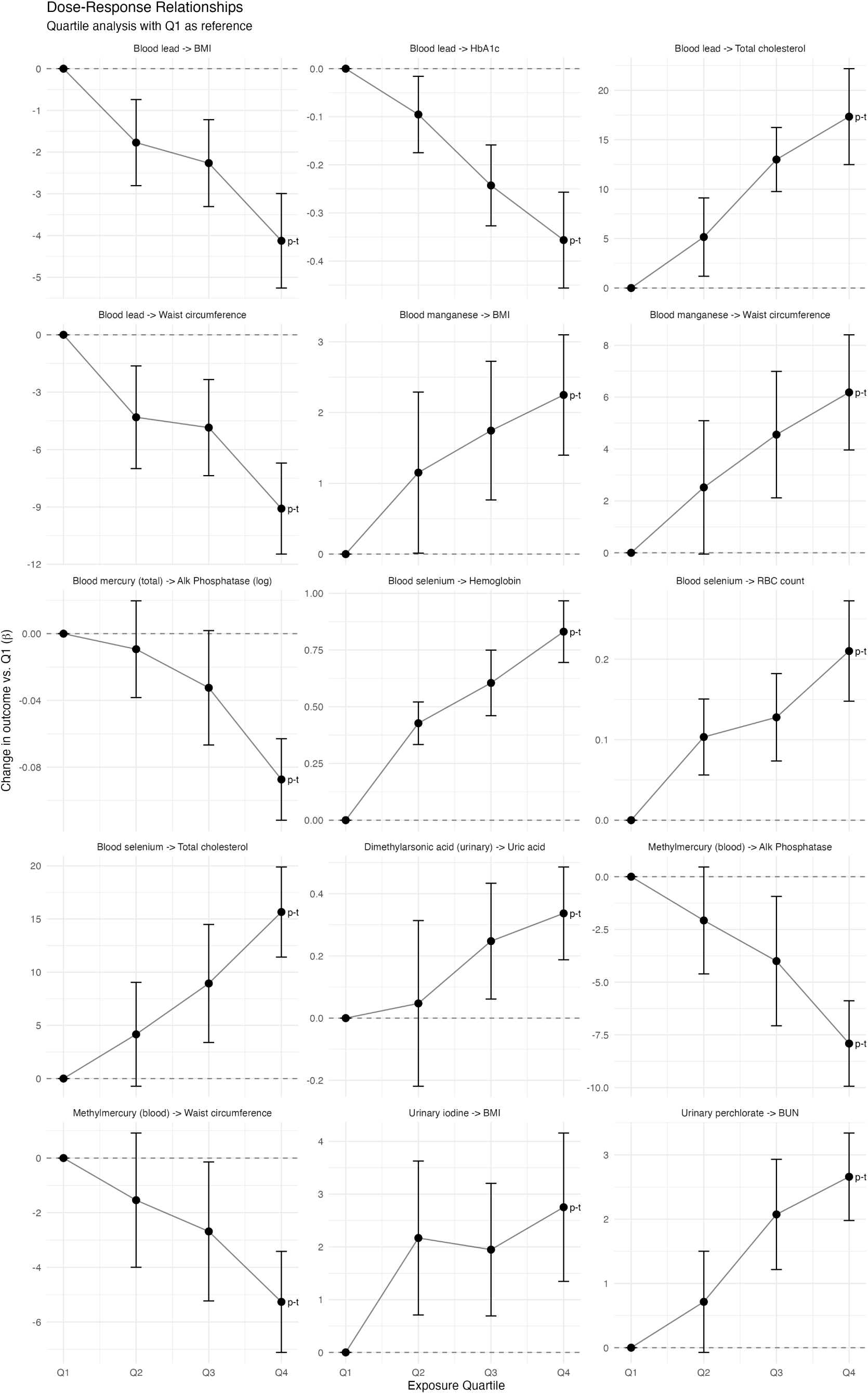
Dose–response curves (quartile analysis) for 15 validated associations. Points represent survey-weighted adjusted mean differences relative to the lowest quartile.

The strongest dose–response gradient was observed for blood lead and total cholesterol: compared to the lowest quartile, mean total cholesterol was 5.2, 13.0, and 17.3 mg/dL higher in the second, third, and fourth quartiles, respectively (p_trend_ = 7.7 × 10^-5^). The single non-monotonic finding was urinary iodine with BMI, where the third quartile effect (*β* = 1.95) was slightly lower than the second (*β* = 2.17), although the overall trend remained significant (p_trend_ = 0.004) and the fourth quartile showed the largest effect (*β* = 2.75).

### 3.4 Sensitivity Analyses

All 15 initially validated findings were classified as robust under the nine primary sensitivity specifications, with concordant effect direction and nominal significance (p < 0.05) in at least 7 of 9 specifications (Figure 4). The median absolute percent change in the *β* coefficient ranged from 0.7% (urinary iodine–BMI) to 13.1% (methylmercury–waist circumference), indicating that point estimates were generally stable across analytic choices. However, analyte-specific sensitivity checks (described below) subsequently identified one association—urinary iodine–BMI—as a dilution artifact, reducing the count of robustly validated findings from 15 to 14.

**Figure 4:**
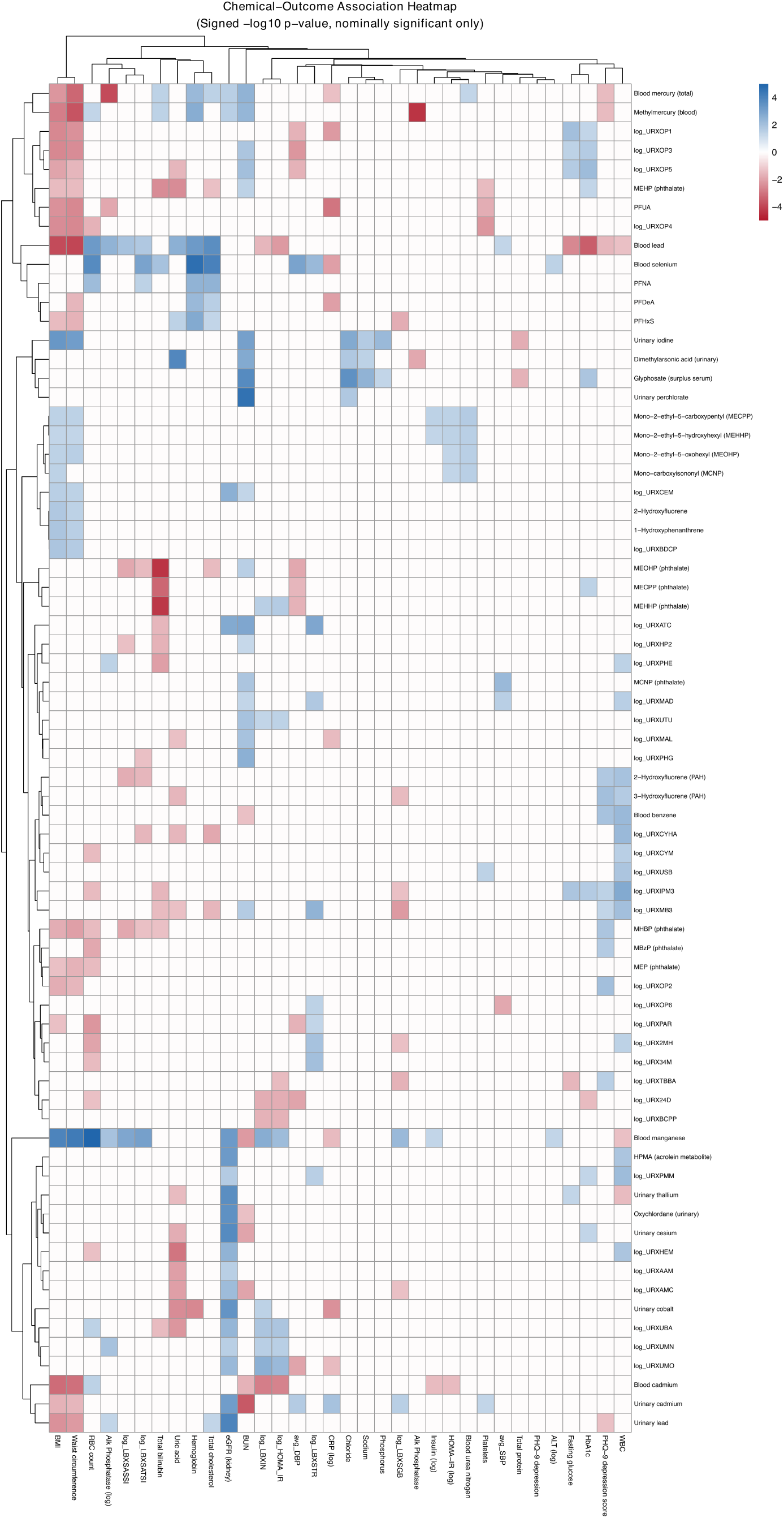
Chemical–outcome association heatmap showing signed log p-value for all nominally significant (p < 0.05) associations. Rows: chemical exposures (clustered by similarity); columns: health outcomes (clustered by domain). Blue = positive; red = negative; intensity reflects significance. Sensitivity results for individual validated findings appear in Figures S1–S15.

Effect estimates held up across sex-stratified and age-stratified subgroups, after exclusion of extreme outliers, with additional adjustment for education, and when substituting serum cotinine (LBXCOT) for the binary smoking indicator. None of the validated associations appeared driven by a single demographic subgroup or by influential outliers. Adding education as an additional covariate—a proxy for socioeconomic position beyond the poverty–income ratio already in the primary model—did not materially change any point estimate, which suggests the primary covariate set adequately captures socioeconomic confounding. Replacing self-reported smoking with continuous cotinine, an objective nicotine biomarker, likewise produced near-identical results; this rules out smoking misclassification as an explanation for any of the observed signals.

Individual sensitivity forest plots for each validated finding are provided in the Supplementary Materials (Figures S1–S15).

#### 3.4.1 Additional sensitivity analyses

Four additional sensitivity specifications targeted specific confounding concerns (Table S5). Adjusting for fish consumption frequency (DBD895, number of fish/shellfish meals in 30 days) did not materially change any of the 15 initially validated findings: all estimates shifted less than 1%, including the methylmercury– waist circumference association (*β* changed from −1.78 to −1.80, 0.8% increase) and both mercury–ALP associations. Physical activity adjustment (18.7% of participants met WHO guidelines for ≥ 150 minutes/week moderate-equivalent activity) similarly produced minimal change: all 15 findings remained significant with effect estimate changes less than 2%. Alcohol consumption adjustment was feasible for 3,143 blood biomarker participants and 1,035 urinary subsample participants with complete data; all 15 findings remained significant after alcohol adjustment, with median absolute effect estimate change of 2.4% (range: −11.6% to +3.2%). For the two HIGH-novelty urinary findings in the alcohol-available subset (n ≈ 1,050), alcohol adjustment produced minimal change: DMA–uric acid *β* = 0.211 (vs. 0.213 without alcohol, −0.9% change) and perchlorate–BUN *β* = 1.39 (vs. 1.42 without alcohol, −2.4% change); both remained highly significant. Effect estimates were predominantly attenuated (12 of 15 findings) rather than amplified, though the magnitude of changes was modest and no finding lost significance, suggesting alcohol is not a major confounder for these associations.

Urinary creatinine adjustment had important implications for the three urinary biomarker associations. Perchlorate–BUN was robust (*β* changed from 1.21 to 1.22, +0.5%, p = 0.001). DMA–uric acid was attenuated by 33% (*β*: 0.202 → 0.135) but remained statistically significant (p = 0.012), suggesting that urinary concentration partially but not fully explains this association. In contrast, the urinary iodine–BMI association was effectively eliminated by creatinine adjustment (*β* changed from 1.18 to 0.40, −66%, p = 0.15), identifying it as a probable artifact of urinary dilution variation rather than a genuine exposure–outcome relationship. This reduces the count of robustly validated findings from 15 to 14.

For the two HIGH-novelty urinary findings (DMA–uric acid and perchlorate–BUN), additional adjustment for dietary protein intake (24-hour recall mean = 79.5 g/day) was conducted because both uric acid and BUN are influenced by protein metabolism (Table S8). This analysis was restricted to participants with complete dietary recall data (N = 1,529 for DMA–uric acid vs. 1,593 in the primary analysis; N = 1,515 for perchlorate–BUN vs. 1,579 in the primary analysis). DMA–uric acid was attenuated by 12% (*β*: 0.202 → 0.178, p = 0.0008), while perchlorate–BUN showed 9% attenuation (*β*: 1.21 → 1.10, p = 0.0002). Both findings remained statistically significant after protein adjustment, indicating that dietary protein intake does not explain these associations.

Sensitivity analysis using the full six-level NHANES RIDRETH3 race/ethnicity classification was conducted for 13 blood biomarker validated findings (Table S7). All 13 findings remained statistically significant (p < 0.0005) with the finer racial/ethnic categorization. Effect estimate changes were uniformly small, ranging from 0.0% (blood lead–HbA1c) to 1.8% (methylmercury–waist circumference and methylmercury–ALP), with a median absolute change of 0.3%. These minimal changes confirm that the three-level race categorization used in the primary analysis did not introduce meaningful confounding bias.

The study was adequately powered to detect small effects: at the FDR-corrected significance level, the minimum detectable *R*^2^ was 1.2% for blood biomarker analyses (effective n = 2,435), 3.6% for urinary subsample analyses (effective n = 790), and 4.2% for surplus serum analyses (effective n = 685). These thresholds are relevant for interpreting null findings (e.g., the PFAS–thyroid screen) but do not validate positive findings, which stand on cross-cycle replication. Full power analysis details are provided in Table S10.

### 3.5 Novelty Assessment

Structured literature review classified the 15 validated findings into four novelty tiers (Table 3):

**Table 3.** Novelty assessment for 15 cross-cycle validated findings (including 1 subsequently identified as artifact).

| Chemical | Outcome | $\beta$ | 95% CI | Novelty | PubMed | |
| --- | --- | --- | --- | --- | --- | --- |
|  |  |  |  |  | Hits | Rationale |
| Methylmercury | Waist circumference<sup>b</sup> | -1.78 | (-2.34, -1.23) | HIGH | 0 | No prior publications; likely confounded by fish consumption |
|  |  |  |  |  | Hits | Rationale |
| DMA<br>(urinary) | Uric acid | 0.20 | (0.15, 0.26) | HIGH | 0 | No publications linking DMA to uric acid; plausible via purine metabolism |
| Urinary perchlorate | BUN | 1.21 | (0.97, 1.45) | HIGH | 2 | Only 2 publications on perchlorate-kidney; BUN in adults not studied |
| Methylmercury | ALP | -2.84 | (-3.47, -2.21) | MODERATE | 1 | Li et al. [12] studied metals-liver, not MeHg-ALP specifically |
| Blood manganese | BMI<sup>b</sup> | 2.59 | (1.87, 3.30) | MODERATE | 2 | Very limited literature; adult population data scarce |
| Blood manganese | Waist circumference<sup>b</sup> | 7.16 | (5.33, 8.99) | MODERATE | 2 | Same sparse literature as Mn-BMI |
|  |  |  |  |  | Hits | Rationale |
| Blood mercury (total) | ALP (log) | -0.04 | (-0.05, -0.03) | MODERATE | 1 | Total Hg-ALP largely unstudied at population level |
| Urinary iodine | BMI<sup>b</sup> | 1.18 | (0.77, 1.58) | ARTIFACT<sup>a</sup> | 8 | Eliminated by creatinine adjustment (dilution artifact) |
| Blood selenium | Hemoglobin | 2.16 | (1.74, 2.58) | LOW-MOD | 4 | Limited but consistent; selenium essential for erythro-poiesis |
| Blood selenium | RBC count | 0.55 | (0.40, 0.70) | LOW-MOD | 4 | Same literature base as selenium-hemoglobin |
| Blood lead | BMI<sup>b</sup> | -1.95 | (-2.51, -1.38) | LOW | 29 | Well-documented; subject to volume dilution bias |
|  |  |  |  |  | Hits | Rationale |
| Blood lead | Waist circumference<sup>b</sup> | -4.35 | (-5.58, -3.13) | LOW | 29 | Same literature as lead-BMI; volume dilution bias |
| Blood lead | HbA1c | -0.19 | (-0.25, -0.13) | LOW | 262 | Extensively studied; inverse at low exposure levels |
| Blood lead | Total cholesterol | 9.10 | (6.55, 11.65) | LOW | 53 | Well-established via oxidative stress pathway |
| Blood selenium | Total cholesterol | 39.59 | (30.02, 49.15) | LOW | 22 | Well-documented across multiple populations |

<sup>a</sup> Urinary iodine–BMI association eliminated after creatinine adjustment ( $\beta$ : 1.18 → 0.40, $p = 0.15$ ); reclassified from MODERATE to ARTIFACT and excluded from robust finding counts.
<sup>b</sup> Models for BMI and waist circumference outcomes omit BMI from the covariate set to avoid collider bias; effect sizes not directly comparable to other findings.

**HIGH novelty (n = 3):** Three associations had no or minimal prior literature. (i) Dimethylarsonic acid (DMA) was positively associated with uric acid—no prior publications specifically examined this arsenic species–uric acid relationship, though broader arsenic–metabolic links have been reported. (ii) Urinary perchlorate was positively associated with BUN, with only two prior publications on perchlorate–kidney function, neither examining BUN in adults. (iii) Methylmercury was inversely associated with waist circumference, a finding with no prior publications but likely confounded by fish consumption.

**MODERATE novelty (n = 4, plus 1 artifact):** Four associations involved chemicals with emerging but limited evidence: methylmercury with ALP, blood manganese with BMI and waist circumference, and total mercury with ALP. A fifth association initially classified as MODERATE—urinary iodine with BMI—was subsequently identified as a probable dilution artifact after creatinine adjustment (see Section 3.4) and is excluded from the robust finding count.

**LOW-MODERATE (n = 2):** Two findings—selenium with hemoglobin and RBC count—had limited but consistent prior literature supporting selenium’s role in erythropoiesis.

**LOW novelty (n = 5):** Five associations were well-established in the literature, including blood lead with BMI, waist circumference, total cholesterol, and HbA1c, and selenium with total cholesterol. These serve as positive controls, confirming that the analytic pipeline recapitulates known biology.

## 4 Discussion

This comprehensive ExWAS of NHANES 2017–2018 systematically screened 2,796 chemical biomarker– health outcome combinations and identified 15 cross-cycle validated associations, of which 14 survived analyte-specific sensitivity checks (urinary creatinine adjustment identified the iodine–BMI association as a dilution artifact), through a rigorous four-stage pipeline: FDR correction, cross-cycle replication, dose– response assessment, and multi-specification sensitivity analysis. Among these, two HIGH-novelty findings (DMA–uric acid, perchlorate–BUN) survived all sensitivity checks, a third HIGH-novelty association (methylmercury–waist circumference) is likely explained by dietary confounding, and four MODERATE- novelty findings represent potentially important contributions to environmental epidemiology.

### 4.1 Novel Findings

#### Dimethylarsonic acid and uric acid

The positive association between urinary DMA (URXUDMA, the primary methylated arsenic metabolite) and serum uric acid (LBXSUA) is, to our knowledge, previously unreported. The observed link aligns with known mechanisms of arsenic-induced oxidative stress: reactive oxygen species generated during arsenic methylation upregulate xanthine oxidase, the terminal enzyme in purine catabolism that converts hypoxanthine to uric acid. Arsenic also impairs renal tubular urate transport, potentially reducing clearance. Prior studies have associated total arsenic with metabolic syndrome components [13–15], but none have isolated DMA—the dominant urinary arsenic species in methylation-competent individuals—as a specific predictor of serum uric acid. Individual variation in arsenic methylation efficiency, driven largely by AS3MT gene polymorphisms [16,17], determines the fraction of total arsenic excreted as DMA. Whether high urinary DMA reflects high arsenic exposure, efficient methylation, or both has implications for interpreting this association; methylation-efficient individuals may generate more DMA-related reactive intermediates, linking genotype to downstream metabolic effects. NHANES does not collect genotype data, so this potential effect modification could not be assessed.

The effect size (*β* = 0.20 mg/dL per log-unit increase) is modest in absolute terms, but given that the interquartile range of log-DMA spans roughly 1.5 units, the implied difference between high and low DMA exposure is approximately 0.30 mg/dL. For context, serum uric acid levels above 6.8 mg/dL define hyperuricemia, which affects approximately 20% of US adults; even small population-level shifts in the uric acid distribution could meaningfully alter gout and cardiovascular risk at the tail. Adjustment for urinary creatinine attenuated the DMA–uric acid estimate by 33% (*β*: 0.202 → 0.135, p = 0.012), indicating that urinary concentration contributes to but does not fully explain the association. The creatinine-adjusted estimate implies an IQR-associated difference of approximately 0.20 mg/dL (0.135 × 1.5)—smaller but still potentially meaningful at the population tail, where even modest rightward shifts in uric acid could push susceptible individuals above clinical thresholds. While this p-value alone would not provide strong evidence in a single-test framework, the convergence of multiple lines of evidence—cross-cycle validation (p = 0.003 in 2015–2016), significant monotonic dose–response, biological plausibility via oxidative stress mechanisms, and persistence after protein intake adjustment—collectively supports a genuine DMA–uric acid relationship.

#### Perchlorate and BUN

Perchlorate is primarily recognized as a thyroid disruptor via competitive inhibition of the sodium-iodide symporter [18,19]. Our finding of a positive perchlorate–BUN association, validated across two NHANES cycles, suggests previously unrecognized renal effects. Only two prior publications have examined perchlorate–kidney function in NHANES: Li et al. [20] focused on eGFR in adults, and Xue et al. [21] studied adolescents. Neither examined BUN directly. The observed *β* of 1.21 mg/dL per log-unit increase in urinary perchlorate (URXUP8) translates to roughly a 2.7 mg/dL difference between the lowest and highest exposure quartiles. Against a normal BUN reference range of 7–20 mg/dL, this represents a shift of approximately 15–20% of the clinical range—large enough to be physiologically relevant, particularly for individuals near the upper boundary. Applied to the roughly 218 million US adults represented by NHANES, a rightward shift of this magnitude in the BUN distribution could push a non-trivial fraction of the population above clinically elevated thresholds, though the precise prevalence impact would depend on the shape of the underlying distribution and the causal nature of the association. Reverse causation cannot be excluded in this cross-sectional design: impaired renal function could reduce perchlorate clearance, increasing urinary perchlorate concentration while simultaneously elevating BUN through diminished urea excretion, thereby creating a spurious positive association. However, a sensitivity analysis adjusting for eGFR showed only 8.5% attenuation of the effect estimate (*β*: 1.21 → 1.11) while remaining highly significant (p = 2.4 × 10^−5^; Table S15), suggesting the association is not primarily driven by renal confounding. BUN levels are also influenced by dietary protein intake and hydration status, neither of which was controlled in our models. If perchlorate exposure correlates with dietary patterns through shared sources (e.g., leafy vegetables, drinking water), the observed association could be partly confounded by these unmeasured factors. Notably, hydration status would likely bias *against* finding a positive association: higher water intake increases urinary perchlorate excretion while simultaneously diluting BUN, so any genuine perchlorate–BUN relationship would be underestimated in individuals with high fluid intake. This consideration strengthens rather than weakens the case for a real association. Notably, adjustment for urinary creatinine did not attenuate this association (*β* changed by 0.5%, p = 0.001), indicating that the perchlorate–BUN signal is not driven by urinary dilution variation.

#### Methylmercury and waist circumference

The inverse association between blood methylmercury (LBXBGM) and waist circumference (BMXWAIST) is statistically robust but almost certainly confounded by fish consumption patterns. Regular fish consumers have both higher methylmercury levels and healthier metabolic profiles [22,23]; adjustment for crude fish frequency (DBD895) and 24-hour dietary recall fish consumption did not attenuate this association, but these variables likely fail to capture the full confounding pathway. Residual dietary confounding remains the most parsimonious explanation.

### 4.2 Moderate-Novelty Findings

Blood manganese (LBXBMN) was positively associated with both BMI and waist circumference, with monotonic dose–response gradients and cross-cycle validation. Manganese is essential for neuronal health and enzymatic function [24], but the evidence linking elevated blood levels to metabolic dysregulation is thin. Smith et al. [25] reported associations between prenatal manganese exposure and childhood metabolic markers; Lo et al. [26] found associations between blood and urinary manganese and metabolic syndrome components in NHANES 2011–2016. Our results add adult population-level evidence from two independent NHANES cycles, with a clear dose–response gradient: participants in the highest LBXBMN quartile had BMI approximately 2.2 kg/m^2^ higher than those in the lowest. Whether elevated manganese drives adiposity through disrupted insulin signaling, or whether adiposity alters manganese metabolism, cannot be resolved cross-sectionally.

Urinary iodine (URXUIO) was positively associated with BMI in both the discovery and validation cycles, but creatinine adjustment eliminated this association (see Section 3.4), identifying it as a probable artifact of urinary dilution rather than genuine iodine exposure. This finding illustrates an important methodological lesson: urinary biomarker associations that survive FDR correction, cross-cycle validation, and multiple demographic sensitivity analyses can still be explained by measurement artifacts when analyte-specific confounders are addressed.

Both methylmercury (LBXBGM) and total mercury (LBXTHG) were inversely associated with ALP. Li et al. [12] examined blood metal mixtures and liver function in NHANES 2011–2018 but focused on mercury– bilirubin relationships rather than the mercury–ALP link we identified. ALP has dual hepatic and osseous origins, and mercury’s inverse association could reflect suppression of osteoblast activity, direct hepato- cyte effects, or—as with the waist circumference finding—confounding by fish intake, since fish consumers may differ from non-consumers in ways that affect ALP. Adjustment for self-reported fish consumption (DBD895) did not attenuate either mercury–ALP association (*β* changed less than 1%), paralleling the methylmercury–waist circumference finding. While the crude fish frequency variable may be insufficient to fully capture dietary confounding, the consistency across methylmercury and total mercury, two NHANES cycles, multiple sensitivity specifications, and fish-adjusted models strengthens the case for a genuine mercury–ALP association.

### 4.3 Established Associations as Positive Controls

Five LOW-novelty findings—blood lead (LBXBPB) with BMI, waist circumference, total cholesterol, and HbA1c, and selenium (LBXBSE) with total cholesterol—serve as positive controls. All five are well-documented in the NHANES literature [27,28]. However, the interpretation of the inverse lead–BMI and lead–waist associations warrants caution. While these findings are consistent with prior NHANES analyses, they are vulnerable to reverse causation via volume dilution: higher body mass dilutes blood lead concentration across a larger blood volume, creating a mechanical inverse association independent of any toxicological mechanism. This bias is one of the most well-documented artifacts in lead epidemiology. Thus, while our pipeline successfully replicates these prior findings, this may reflect consistent measurement of the same bias rather than validation of a true biological relationship. The lead–BMI/waist findings demonstrate that our pipeline detects associations previously reported in NHANES, but they cannot serve as unambiguous positive controls in the sense of confirming true biological effects. Lead–cholesterol tracks with lead’s effects on lipid peroxidation, while the inverse lead–HbA1c association—seemingly paradoxical—has been reported at the low environmental exposure levels typical of the contemporary US population and may reflect non-linear dynamics or healthy-survivor bias. The lead–HbA1c finding showed minimal change (−1.2%) with quadratic age adjustment (Table S13), suggesting the association is not explained by non-linear age confounding despite prior reports of age-dependent lead–glycemic relationships. Selenium–cholesterol is one of the most replicated ExWAS findings across NHANES cycles and international cohorts. That these known associations emerged from the same pipeline that identified novel signals provides confidence that the framework has adequate sensitivity at population-level exposure concentrations, while the caveats around lead–anthropometric associations illustrate the limitations of cross-sectional biomarker studies.

### 4.4 Strengths

A hypothesis-free design that screens a broad chemical–outcome space is inherently less susceptible to publication bias than targeted studies, and increases the chances of detecting overlooked associations. Unlike prior single-stage screens, we enforced a strict validation architecture: every finding had to survive FDR correction across 2,796 tests, replicate in an independent NHANES cycle, show a monotonic dose–response gradient, and remain robust across nine sensitivity specifications. This four-stage filter is, to our knowledge, the most stringent validation framework applied in an ExWAS to date. Standardized CDC laboratory methods across both discovery (2017–2018) and validation (2015–2016) cycles ensure measurement comparability. The breadth of the screen—diverse chemical classes and outcome domains analyzed within a single framework—also enables comparison of relative effect sizes under identical analytic conditions. Direct magnitude comparisons require caution when anthropometric outcomes are involved, however, as those models omit BMI from the covariate set to avoid collider bias (see Table 3, footnote b).

### 4.5 Limitations

Several limitations bear on interpretation. The cross-sectional design precludes causal inference. Reverse causation is a particular concern for biomarkers influenced by disease status—eGFR alters urinary metal excretion independently of true exposure. The inverse lead–BMI and lead–waist circumference associations, while cross-cycle validated, are particularly vulnerable to reverse causation via volume dilution: higher body mass expands the blood volume over which lead distributes, mechanically lowering measured blood lead concentration independent of any toxicological effect. This phenomenon is well-documented in lead epidemiology [29] and represents a limitation of blood lead as an exposure biomarker in cross-sectional anthropometric studies. While we classify these associations as “LOW novelty” positive controls because they replicate prior NHANES findings, this consistency may partly reflect consistent measurement of the same bias rather than a true biological relationship.

Residual confounding by diet, physical activity, occupational exposures, and medication use cannot be excluded despite adjustment for six covariates. Adjustment for self-reported fish/shellfish consumption frequency (DBD895) did not attenuate the mercury–waist circumference or mercury–ALP findings, but this single-item variable (number of fish meals in 30 days) is a poor proxy for fish consumption patterns that drive methylmercury exposure. Mercury bioaccumulates differentially across fish species (e.g., tuna vs. tilapia), and the crude frequency measure cannot capture this variability. Additional adjustment using 24-hour dietary recall fish consumption (grams consumed on the recall day, derived from individual food codes) also showed no attenuation of the mercury findings (|Δ*β*| < 1% for all three associations; Table S12). This creates a puzzle: if fish consumption truly explains the methylmercury–waist association (through correlated healthy dietary patterns), we would expect fish adjustment to attenuate the effect—yet neither self-reported frequency nor 24-hour recall adjustment did so. This persistence could indicate a genuine direct effect of methylmercury on adiposity, or it may simply reflect that available fish consumption measures are too crude to capture the relevant dietary patterns. A single 24-hour recall inadequately captures habitual fish intake given methylmercury’s long biological half-life (weeks to months), and species-specific consumption data would be needed to properly distinguish toxicological from dietary effects. We therefore cannot definitively adjudicate between direct mercury toxicity and residual dietary confounding as explanations for this association. Alcohol consumption adjustment was feasible only for a subset of participants (n = 3,143 for blood biomarkers, n = 1,035 for urinary subsample) due to NHANES ALQ_J skip patterns; while all findings remained significant in this reduced sample, the smaller effective sample sizes limit the precision of these estimates.

The three-level race/ethnicity classification (Non-Hispanic White, Non-Hispanic Black, Other) was adopted to ensure model stability across the many exposure–outcome combinations, particularly in subsample analyses where finer categories would produce sparse cells. However, collapsing Mexican American, Other Hispanic, Non-Hispanic Asian, and Other/Multiracial into a single “Other” category represents a substantial loss of granularity that could mask important effect modification—particularly for arsenic findings, where methylation efficiency varies by ancestry, and lead findings, where exposure disparities are well-documented by race/ethnicity. The six-level sensitivity analysis (Table S7) showed that all blood biomarker findings remained significant with finer categorization, with effect estimate changes uniformly less than 2%, which is reassuring but does not fully address the potential for masked heterogeneity.

The primary model used only linear age, without a quadratic term to capture non-linear age effects. For findings like lead–HbA1c, where non-linear relationships have been reported in the literature, this specification may incompletely adjust for age-related confounding. A sensitivity analysis including quadratic age (age²) showed that all 15 validated findings maintained direction and significance (Table S13), with effect estimate changes from −11% to +14%, indicating that non-linear age effects do not explain the primary findings. Models using surplus serum (WTSSBJ2Y) or urinary subsample (WTSA2YR) weights had low residual degrees of freedom (df = 7–9). This arises because NHANES uses only 15 primary sampling units across 2 variance strata, yielding a maximum of 13 residual df for variance estimation before accounting for model parameters; subsample analyses with fewer non-empty strata reduce this further. Critically, low df makes inference *more* conservative, not less: the t-distribution with df = 8 has heavier tails than the normal, producing wider confidence intervals and larger p-values for any given effect size. Any urinary or surplus serum finding that achieved significance despite this penalty reflects a stronger underlying signal than its nominal p-value suggests. This conservative bias also motivated our use of a parsimonious six-covariate model rather than a more elaborate specification that would consume additional degrees of freedom.

The novelty assessment relied on PubMed keyword searches, which may miss relevant studies indexed under different MeSH terms or published in non-English journals. Supplementary MeSH-based searches (Table S14) retrieved 36 articles for DMA–uric acid, 72 for perchlorate–BUN, and 51 for methylmercury–waist circumference; however, manual review confirmed these articles address the broader chemical classes (e.g., arsenic, general kidney toxicology) rather than the specific exposure–outcome pairs we identified. The novelty classifications therefore remain appropriate, but Embase searches would further strengthen these claims. The sequential adaptive design—where the expanded and novelty screens were conducted because earlier screens yielded null results—means that the global FDR correction is optimistic; the effective multiple testing burden exceeds the nominal 2,796 tests. Cross-cycle validation provides the primary protection against false positives, and the within-round FDR analysis (Table S6) shows that results are robust to this concern. Finally, while the LOD/√2 substitution is standard practice, it can introduce bias when non-detect rates are high—though as noted, none of the validated chemicals approached the 70% exclusion threshold.

### 4.6 Comparison with Prior ExWAS Studies

Patel et al. [1] screened 266 environmental factors against type 2 diabetes in the original ExWAS, identifying heptachlor epoxide and gamma-tocopherol as top hits. Subsequent studies have examined serum lipids, liver enzymes, and kidney function [11,30], but most tested fewer than 500 exposure–outcome pairs and relied on single-cycle analyses without systematic validation. Our analysis is substantially larger (2,796 tests across seven clinical domains) and, to our knowledge, the first ExWAS to apply all four validation stages—FDR, cross-cycle replication, dose–response, and multi-specification sensitivity—within a single study. That 71% of testable associations replicated across independent NHANES cycles is reassuring: it suggests that most FDR-corrected ExWAS findings in NHANES reflect genuine signals rather than chance patterns. A caveat is that consecutive NHANES cycles share not only standardized protocols but potentially similar confounding structures—both cycles sampled from the same underlying US population with similar dietary patterns, environmental regulations, and socioeconomic gradients—so replication may partly reflect consistent capture of the same confounded associations rather than purely independent validation. Temporal gaps of 10+ years, or replication in non-US cohorts, would provide stronger tests.

The failure of several discovery-phase findings to validate across cycles is instructive. Most strikingly, blood manganese–RBC count was the single Bonferroni-significant result in 2017–2018 yet showed a reversed direction in 2015–2016, underscoring why cross-cycle validation—not p-value magnitude—is the appropriate criterion for advancing ExWAS findings. Urinary biomarker concentrations are influenced by renal function itself, creating a circularity problem in cross-sectional studies: impaired kidney function reduces urinary excretion, altering biomarker concentrations independently of true exposure. This reverse causation bias is a well-recognized limitation for urinary metals in kidney outcome studies and likely explains why the cesium–eGFR, thallium–eGFR, lead–eGFR, and cobalt–eGFR associations were significant in 2017–2018 but did not replicate in 2015–2016.

The null PFAS–thyroid result merits comment. We screened 60 PFAS–thyroid combinations (6 PFAS biomarkers × 10 thyroid markers) and found no FDR-significant associations. This is consistent with the mixed epidemiological literature on PFAS and thyroid function [31], where associations have been inconsistent across studies, often null in general population samples, and more pronounced in occupationally exposed cohorts or vulnerable subpopulations. The power analysis (Table S10) confirms that our blood biomarker sample (effective n ≈ 2,435) had 80% power to detect effects explaining as little as 1.2% of outcome variance at the FDR-corrected threshold—comparable to or smaller than the effect sizes reported in positive PFAS–thyroid studies. The absence of signal in this well-powered screen suggests that if PFAS–thyroid associations exist at environmental exposure levels in the general US adult population, they are likely smaller than commonly assumed or confined to susceptible subgroups not captured by population-average analyses.

### 4.7 Implications

The three HIGH-novelty findings—DMA with uric acid, perchlorate with BUN, and methylmercury with waist circumference—warrant distinct follow-up approaches. The DMA–uric acid association is the most promising for targeted investigation, as it lacks the dietary confounding that complicates the mercury findings and—despite 33% attenuation after creatinine adjustment—remains statistically significant, supporting a genuine pathway from arsenic methylation to purine metabolism. Prospective cohort studies with serial biomarker measurements could establish temporality and dose–response at the individual level. The perchlorate–BUN finding, if confirmed prospectively, could expand the recognized health effects of perchlorate exposure beyond thyroid disruption to include renal function, with implications for drinking water standards. Two additional associations—glyphosate with BUN and glyphosate with chloride—reached FDR significance in the discovery phase but could not be validated because glyphosate was not measured in the 2015– 2016 surplus serum panel. These findings remain candidates for future validation when glyphosate biomonitoring becomes available in newer NHANES cycles. Linkage to the National Death Index—available for NHANES participants through public-use mortality files—would allow testing whether cross-sectional chemical–biomarker associations predict mortality outcomes; this represents a planned future extension of the current analysis.

At a methodological level, the fact that this pipeline simultaneously recovered well-established associations (lead–cholesterol, selenium–cholesterol) and surfaced genuinely new signals (DMA–uric acid, perchlorate– BUN) suggests that multi-stage ExWAS can be both sensitive and specific. The key is the validation architecture: any single-stage screen of 2,796 tests will produce false positives, but requiring FDR significance, cross-cycle replication, dose–response confirmation, and sensitivity robustness filters these out effectively. We would encourage future ExWAS studies to adopt similar multi-stage frameworks, particularly cross-cycle validation, which proved to be the most informative filter in this analysis.

## 5 Conclusions

In this multi-stage ExWAS of 2,796 chemical biomarker–health outcome associations in NHANES 2017– 2018, we identified 15 cross-cycle validated findings, of which 14 proved robust after analyte-specific sensitivity checks (urinary creatinine adjustment identified iodine–BMI as a dilution artifact). Of three HIGH-novelty associations, two—dimethylarsonic acid with uric acid and perchlorate with blood urea nitrogen— survived all sensitivity checks including dietary confounding assessment and represent genuinely novel findings warranting prospective investigation; the third (methylmercury–waist circumference) is likely explained by fish consumption patterns. The identification of the iodine–BMI association as a dilution artifact demonstrates the value of analyte-specific sensitivity checks beyond standard demographic robustness analyses. These results demonstrate the value of systematic, multi-stage ExWAS for generating robust hypotheses from population biomonitoring data.

## Data Availability

All data produced in the present work are available online at https://github.com/hayden-farquhar/NHANES-EXWAS.

## Abbreviations

ALP: alkaline phosphatase
BMI: body mass index
BUN: blood urea nitrogen
DMA: dimethylarsonic acid
eGFR: estimated glomerular filtration rate
ExWAS: environment-wide association study
FDR: false discovery rate
HbA1c: glycated hemoglobin
LOD: limit of detection
NHANES: National Health and Nutrition Examination Survey
PIR: poverty–income ratio
RBC: red blood cell

## 6 Ethics Statement

This study used publicly available, de-identified data from the National Health and Nutrition Examination Survey (NHANES). No ethics approval was required as NHANES data are collected under existing IRB protocols at the National Center for Health Statistics, and all data are publicly available for research use. All participants provided written informed consent to the original NHANES protocol.

## 7 Data and Code Availability

This study is reported in accordance with the STROBE guidelines [32] for cross-sectional studies; a completed STROBE checklist is provided in Table S11.

All data used in this study are publicly available from the CDC NHANES website (https://wwwn.cdc.gov/nchs/nhanes/). Analysis code is available at https://github.com/hayden-farquhar/NHANES-EXWAS. NHANES data are downloaded programmatically via the nhanesA R package within the provided scripts.

## 10 Author Contributions (CRediT)

**Hayden Farquhar:** Conceptualization, Methodology, Software, Formal Analysis, Investigation, Data Curation, Writing – Original Draft, Writing – Review & Editing, Visualization.

