## Supplementary materials for "Environment-Wide Association Study of Chemical Biomarkers and Health Outcomes in NHANES 2017–2018: Discovery, Validation, and Dose–Response Analysis"

Hayden Farquhar, MBBS, MPHTM

February 2026

#### 1 NHANES Variable Name Glossary

LBXBGM, blood methylmercury; LBXBMN, blood manganese; LBXBPB, blood lead; LBXBSE, blood selenium; LBXCOT, serum cotinine; LBXRBCSI, RBC count; LBXSAPSI, serum alkaline phosphatase; LBXSUA, serum uric acid; LBXTHG, total blood mercury; BMXWAIST, waist circumference; URXUDMA, urinary dimethylarsonic acid; URXUIO, urinary iodine; URXUP8, urinary perchlorate. Survey weights: WTMEC2YR (MEC exam, blood biomarkers), WTSA2YR (urinary subsample A), WTSSBJ2Y (surplus serum).

### 2 Table S1. All 26 FDR-Significant Associations from the Discovery Phase

Survey-weighted linear regression of log-transformed chemical biomarker on health outcome, adjusted for age, sex, race/ethnicity, poverty–income ratio, BMI (when not the outcome), and smoking status. 95% CIs calculated as $\beta \pm 1.96 \times$ SE.

| Chemical | Outcome | Class | $\beta$ | 95% CI | SE | P-value | FDR | N |
|---|---|---|---|---|---|---|---|---|
| Blood manganese | RBC count | Heavy metals | 0.23 | (0.19, 0.27) | 0.020 | 9.8e-06 | 0.024 | 4,873 |
| Blood selenium | Hemoglobin | Heavy metals | 2.16 | (1.74, 2.58) | 0.215 | 2.0e-05 | 0.024 | 4,873 |
| Urinary perchlorate | BUN | Urinary elements | 1.21 | (0.97, 1.45) | 0.124 | 2.5e-05 | 0.024 | 1,579 |
| MEOHP (phthalate) | Total bilirubin | Phthalates | -0.06 | (-0.07, -0.05) | 0.006 | 4.0e-05 | 0.024 | 1,580 |
| Methylmercury | Alk Phosphatase | Heavy metals | -2.84 | (-3.47, -2.21) | 0.321 | 4.8e-05 | 0.024 | 4,834 |
| Blood manganese | Waist circumference | Heavy metals | 7.16 | (5.33, 8.99) | 0.934 | 5.9e-05 | 0.024 | 4,693 |
| MEHHP (phthalate) | Total bilirubin | Phthalates | -0.06 | (-0.08, -0.05) | 0.006 | 6.1e-05 | 0.024 | 1,580 |
| Blood selenium | Total cholesterol | Heavy metals | 39.59 | (30.02, 49.15) | 4.880 | 8.3e-05 | 0.029 | 4,855 |

| Chemical | Outcome | Class | $\beta$ | 95% CI | SE | P-value | FDR | N |
|---|---|---|---|---|---|---|---|---|
| Blood manganese | BMI | Heavy metals | 2.59 | (1.87, 3.30) | 0.367 | 1.1e-04 | 0.030 | 4,876 |
| Blood lead | Waist circumference | Heavy metals | -4.35 | (-5.58, -3.13) | 0.624 | 1.2e-04 | 0.030 | 4,693 |
| Urinary lead | eGFR | Urinary metals | 6.55 | (5.09, 8.01) | 0.746 | 1.2e-04 | 0.030 | 1,589 |
| Blood lead | BMI | Heavy metals | -1.95 | (-2.51, -1.38) | 0.289 | 1.5e-04 | 0.030 | 4,876 |
| Blood mercury (total) | Alk Phosphatase (log) | Heavy metals | -0.04 | (-0.05, -0.03) | 0.005 | 1.5e-04 | 0.030 | 4,833 |
| DMA (urinary) | Uric acid | Urinary metals | 0.20 | (0.15, 0.26) | 0.027 | 1.5e-04 | 0.030 | 1,593 |
| Urinary cesium | eGFR | Urinary metals | 9.78 | (7.48, 12.08) | 1.174 | 1.6e-04 | 0.030 | 1,589 |
| Glyphosate (serum) | BUN | Surplus serum | 1.22 | (0.89, 1.55) | 0.170 | 1.8e-04 | 0.031 | 1,371 |
| Blood selenium | RBC count | Heavy metals | 0.55 | (0.40, 0.70) | 0.077 | 1.9e-04 | 0.031 | 4,873 |
| Blood lead | Total cholesterol | Heavy metals | 9.10 | (6.55, 11.65) | 1.300 | 2.1e-04 | 0.032 | 4,855 |
| Urinary thallium | eGFR | Urinary metals | 7.68 | (5.78, 9.59) | 0.970 | 2.2e-04 | 0.032 | 1,589 |
| Methylmercury | Waist circumference | Heavy metals | -1.78 | (-2.34, -1.23) | 0.282 | 2.3e-04 | 0.032 | 4,693 |

| Chemical | Outcome | Class | $\beta$ | 95% CI | SE | P-value | FDR | N |
|---|---|---|---|---|---|---|---|---|
| Oxychlordane | eGFR | VOC metabo-lites | 3.23 | (2.39, 4.06) | 0.428 | 2.8e-04 | 0.036 | 1,589 |
| Urinary cadmium | BUN | Urinary metals | -1.02 | (-1.29, -0.76) | 0.137 | 3.0e-04 | 0.036 | 1,589 |
| Glyphosate (serum) | Chloride | Surplus serum | 0.46 | (0.32, 0.59) | 0.069 | 3.0e-04 | 0.036 | 1,374 |
| Blood lead | HbA1c | Heavy metals | -0.19 | (-0.25, -0.13) | 0.029 | 3.4e-04 | 0.039 | 4,874 |
| Urinary cobalt | eGFR | Urinary metals | 3.75 | (2.74, 4.76) | 0.516 | 3.4e-04 | 0.039 | 1,589 |
| Urinary iodine | BMI | Urinary elements | 1.18 | (0.78, 1.58) | 0.205 | 4.4e-04 | 0.047 | 1,600 |

### 3 Table S2. Dose–Response Quartile Analysis for 15 Initially Validated Findings

Survey-weighted adjusted mean differences in outcome relative to the lowest exposure quartile (Q1, reference). $P_{trend}$ from linear contrast across quartile midpoints. Note: Urinary iodine–BMI was subsequently identified as a dilution artifact after creatinine adjustment (see Table S5); the final count of robustly validated findings is 14.

| Chemical | Outcome | Q2 $\beta$ | Q3 $\beta$ | Q4 $\beta$ | $P_{trend}$ | Monotonic | N |
|---|---|---|---|---|---|---|---|
| Blood selenium | Hemoglobin | 0.43 | 0.60 | 0.83 | 1.1e-05 | Yes | 4,873 |
| Urinary perchlorate | BUN | 0.71 | 2.07 | 2.66 | 6.3e-05 | Yes | 1,579 |
| Methylmercury | Alk Phosphatase | -2.07 | -4.00 | -7.91 | 1.7e-04 | Yes | 4,834 |
| Blood manganese | Waist circumference | 2.52 | 4.55 | 6.18 | 1.2e-04 | Yes | 4,693 |
| Blood selenium | Total cholesterol | 4.16 | 8.94 | 15.66 | 1.3e-04 | Yes | 4,855 |
| Blood manganese | BMI | 1.15 | 1.74 | 2.25 | 2.0e-04 | Yes | 4,876 |
| Blood lead | Waist circumference | -4.31 | -4.85 | -9.09 | 1.5e-04 | Yes | 4,693 |
| Blood lead | BMI | -1.77 | -2.26 | -4.13 | 1.5e-04 | Yes | 4,876 |

| Chemical | Outcome | Q2 $\beta$ | Q3 $\beta$ | Q4 $\beta$ | $P_{trend}$ | Monotonic | N |
|---|---|---|---|---|---|---|---|
| Blood mercury (total) | Alk Phosphatase (log) | -0.01 | -0.03 | -0.09 | 1.7e-04 | Yes | 4,833 |
| DMA (urinary) | Uric acid | 0.05 | 0.25 | 0.34 | 4.7e-04 | Yes | 1,593 |
| Blood selenium | RBC count | 0.10 | 0.13 | 0.21 | 3.4e-04 | Yes | 4,873 |
| Blood lead | Total cholesterol | 5.16 | 13.00 | 17.33 | 7.7e-05 | Yes | 4,855 |
| Methylmercury | Waist circumference | -1.54 | -2.68 | -5.26 | 3.2e-04 | Yes | 4,693 |
| Blood lead | HbA1c | -0.10 | -0.24 | -0.36 | 2.4e-04 | Yes | 4,874 |
| Urinary iodine | BMI | 2.17 | 1.95 | 2.75 | 4.3e-03 | No | 1,600 |

#### 4 Table S3. Sensitivity Analysis Summary for 15 Initially Validated Findings

Each finding was tested under 9 specifications: (1) primary model, (2) females only, (3) males only, (4) age $< 50$, (5) age $\geq 50$, (6) excluding outliers > 99th percentile, (7) adjusting for education, (8) cotinine instead of binary smoking, and (9) adults aged 20+ only. A finding is "robust" if $\geq 7$ of 9 specifications show concordant direction and $p < 0.05$. Note: Urinary iodine–BMI was subsequently identified as a dilution artifact after creatinine adjustment (see Table S5); the final count of robustly validated findings is 14.

| Chemical | Outcome | Dir. Match | Sig. (p<0.05) | Median % $\Delta\beta$ | Robust |
|---|---|---|---|---|---|
| Blood manganese | BMI | 9/9 | 9/9 | 5.8% | Yes |
| Blood manganese | Waist circumference | 9/9 | 9/9 | 4.1% | Yes |
| Blood lead | BMI | 9/9 | 9/9 | 11.6% | Yes |
| Blood lead | Waist circumference | 9/9 | 9/9 | 11.0% | Yes |
| Urinary iodine | BMI | 9/9 | 9/9 | 0.7% | Yes |
| Urinary perchlorate | BUN | 9/9 | 9/9 | 5.5% | Yes |
| Methylmercury | Alk Phosphatase | 9/9 | 8/9 | 3.1% | Yes |
| Blood lead | HbA1c | 9/9 | 8/9 | 10.5% | Yes |
| Blood lead | Total cholesterol | 9/9 | 8/9 | 6.2% | Yes |
| Blood selenium | Hemoglobin | 9/9 | 8/9 | 8.4% | Yes |
| Blood selenium | RBC count | 9/9 | 8/9 | 2.1% | Yes |
| Blood selenium | Total cholesterol | 9/9 | 8/9 | 2.9% | Yes |

| Chemical | Outcome | Dir. Match | Sig. (p<0.05) | Median % $\Delta\beta$ | Robust |
|---|---|---|---|---|---|
| Blood mercury (total) | Alk Phosphatase (log) | 9/9 | 8/9 | 2.8% | Yes |
| DMA (urinary) | Uric acid | 9/9 | 8/9 | 1.4% | Yes |
| Methylmercury | Waist circumference | 9/9 | 7/9 | 13.1% | Yes |

### 5 Table S4. Detailed Sensitivity Results for HIGH-Novelty Findings

Effect estimates ($\beta$), standard errors, and p-values for each of the 9 sensitivity specifications for the three HIGH-novelty findings.

#### 5.1 DMA (urinary) – Uric acid

| Specification | $\beta$ | SE | P-value | N | $\Delta\beta$ (%) |
|---|---|---|---|---|---|
| Primary model | 0.202 | 0.027 | 1.5e-04 | 1,593 | – |
| Females only | 0.191 | 0.042 | 0.002 | 813 | -5.4 |
| Males only | 0.227 | 0.077 | 0.018 | 780 | +12.4 |
| Age < 50 | 0.204 | 0.055 | 0.008 | 697 | +1.4 |
| Age $\geq$ 50 | 0.179 | 0.084 | 0.070 | 896 | -11.4 |
| Excl. outliers (>P99) | 0.220 | 0.032 | 2.4e-04 | 1,575 | +9.1 |
| Adjust for education | 0.203 | 0.030 | 0.021 | 1,593 | +0.8 |
| Cotinine for smoking | 0.202 | 0.028 | 1.6e-04 | 1,593 | +0.1 |
| Adults 20+ only | 0.202 | 0.027 | 1.5e-04 | 1,593 | – |

#### 5.2 Urinary perchlorate – BUN

| Specification | $\beta$ | SE | P-value | N | $\Delta\beta$ (%) |
|---|---|---|---|---|---|
| Primary model | 1.211 | 0.124 | 2.5e-05 | 1,579 | – |
| Females only | 1.444 | 0.229 | 2.3e-04 | 803 | +19.3 |
| Males only | 0.870 | 0.233 | 0.006 | 776 | -28.1 |
| Age < 50 | 0.965 | 0.161 | 5.4e-04 | 692 | -20.3 |
| Age $\geq$ 50 | 1.382 | 0.238 | 6.6e-04 | 887 | +14.2 |
| Excl. outliers (>P99) | 1.277 | 0.134 | 3.0e-05 | 1,563 | +5.5 |
| Adjust for education | 1.209 | 0.122 | 0.002 | 1,579 | -0.1 |
| Cotinine for smoking | 1.210 | 0.128 | 3.0e-05 | 1,579 | 0.0 |
| Adults 20+ only | 1.211 | 0.124 | 2.5e-05 | 1,579 | – |

#### 5.3 Methylmercury – Waist circumference

| Specification | $\beta$ | SE | P-value | N | $\Delta\beta$ (%) |
|---|---|---|---|---|---|
| Primary model | -1.783 | 0.282 | 2.3e-04 | 4,693 | – |
| Females only | -2.868 | 0.556 | 5.9e-04 | 2,413 | +60.9 |
| Males only | -0.909 | 0.403 | 0.050 | 2,280 | -49.0 |
| Age < 50 | -2.474 | 0.458 | 6.5e-04 | 2,121 | +38.8 |
| Age $\geq$ 50 | -1.166 | 0.551 | 0.067 | 2,572 | -34.6 |
| Excl. outliers (>P99) | -1.886 | 0.297 | 2.2e-04 | 4,643 | +5.8 |
| Adjust for education | -1.549 | 0.330 | 0.042 | 4,693 | -13.1 |
| Cotinine for smoking | -1.788 | 0.284 | 2.3e-04 | 4,693 | +0.3 |
| Adults 20+ only | -1.783 | 0.282 | 2.3e-04 | 4,693 | – |

### 6 Table S5. Additional Sensitivity Analyses

Four additional sensitivity specifications were applied to assess specific confounding concerns. Fish consumption frequency (DBD895, number of fish/shellfish meals in 30 days) and physical activity (PAQ_J) were tested for all 15 initially validated findings. Alcohol consumption (ALQ_J) was assessed in the subset of participants with complete alcohol data (n = 3,143 for blood biomarkers, n = 1,035 for urinary subsample); all 15 findings remained significant with median absolute effect change of 2.4%. Urinary creatinine adjustment (log-transformed) was applied to the three urinary biomarker associations to address dilution variation; this analysis identified urinary iodine–BMI as a dilution artifact (see main text Section 3.4).

#### 6.1 Fish consumption and physical activity adjustment

Adjustment for self-reported fish consumption frequency (DBD895, number of fish/shellfish meals in 30 days) did not materially change any of the 15 initially validated findings (all $|\Delta\beta| < 1\%$). For mercury findings specifically, an additional sensitivity analysis using 24-hour dietary recall fish consumption (grams consumed) is presented in Table S12, which also showed minimal change ($|\Delta\beta| < 1\%$). Physical activity adjustment likewise produced no change ($|\Delta\beta| < 0.01\%$ for all findings). These results are consistent across all 15 associations and indicate that the primary findings are not confounded by fish intake frequency or recreational physical activity level.

#### 6.2 Urinary creatinine adjustment

| Chemical | Outcome | $\beta$ (primary) | $\beta$ (creatinine-adj) | $\Delta\beta$ (%) | P (creatinine-adj) | Interpretation |
|---|---|---|---|---|---|---|
| Urinary perchlorate | BUN | 1.211 | 1.217 | +0.5% | 0.001 | Robust; not driven by dilution |

| Chemical | Outcome | $\beta$ (primary) | $\beta$ (creatinine-adj) | $\Delta\beta$ (%) | P (creatinine-adj) | Interpretation |
|---|---|---|---|---|---|---|
| DMA (urinary) | Uric acid | 0.202 | 0.135 | -32.9% | 0.012 | Attenuated but significant; partial dilution contribution |
| Urinary iodine | BMI | 1.177 | 0.402 | -65.9% | 0.153 | **Eliminated**; probable dilution artifact |

The perchlorate–BUN association was unchanged by creatinine adjustment, confirming that urinary dilution variation does not explain this signal. The DMA–uric acid association was attenuated by approximately one-third but remained statistically significant, suggesting that urinary concentration contributes to but does not fully account for the observed relationship. The urinary iodine–BMI association was eliminated after creatinine correction, indicating that the original signal reflected systematic differences in urine concentration correlated with body size rather than genuine iodine exposure effects.

### 7 Figures S1–S15. Individual Sensitivity Forest Plots

Forest plots showing the effect estimate ($\beta$) and 95% confidence interval for each of the 9 sensitivity specifications. The vertical dashed line marks $\beta = 0$ (null). Findings are ordered by novelty tier.

#### 7.1 HIGH Novelty

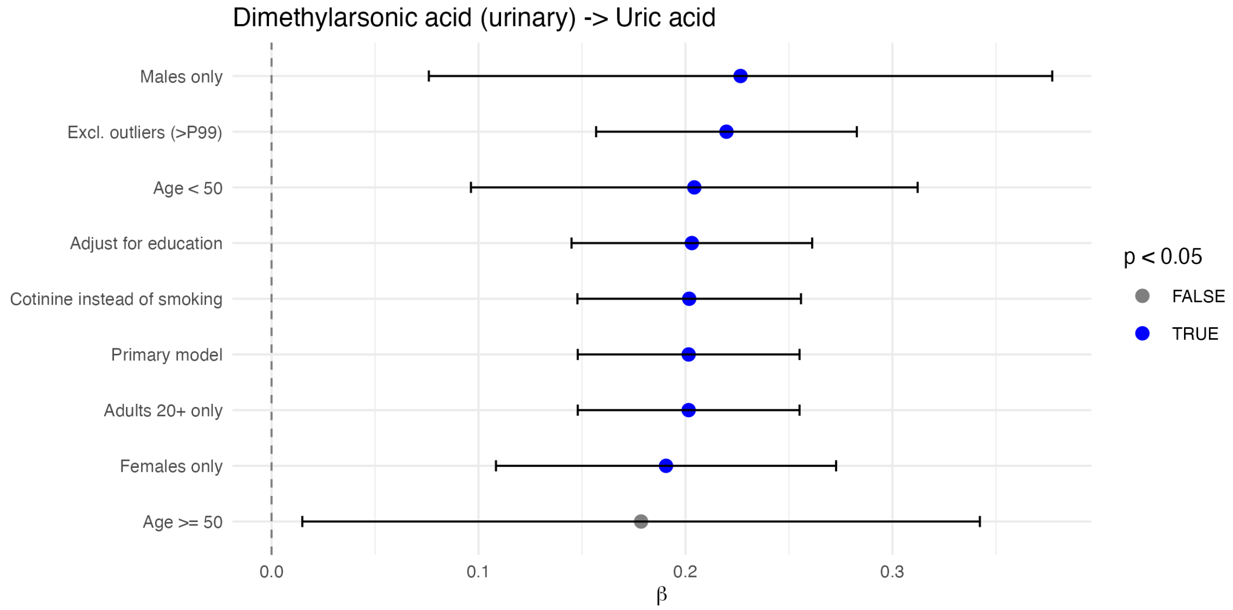


Figure S1: Sensitivity analysis: DMA (urinary) – Uric acid.

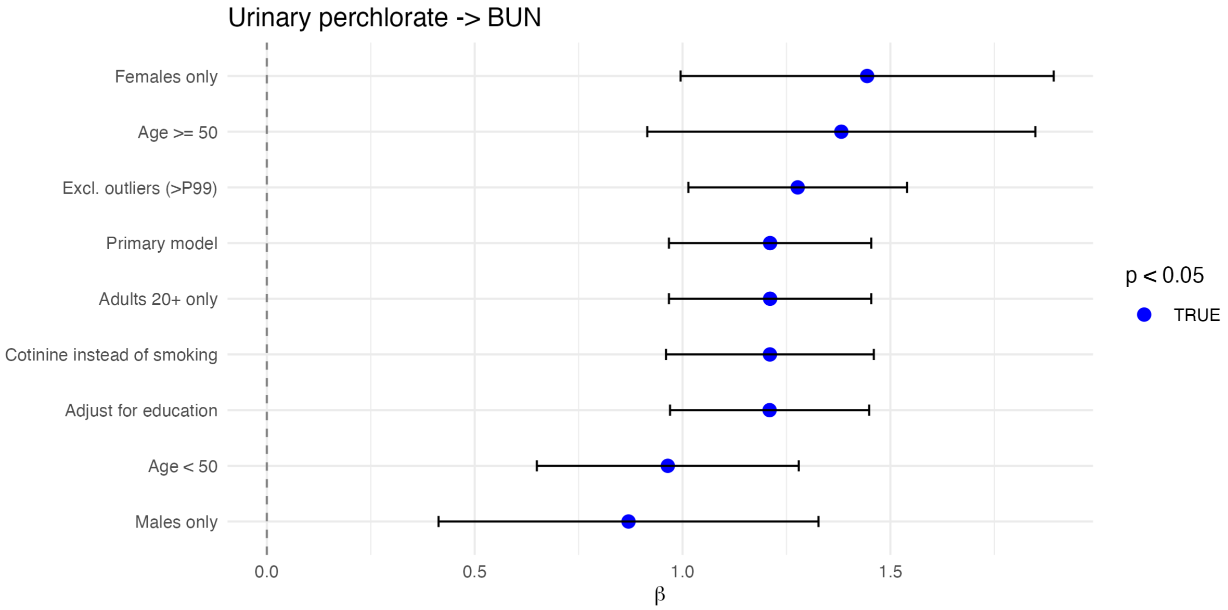


Figure S2: Sensitivity analysis: Urinary perchlorate – BUN.

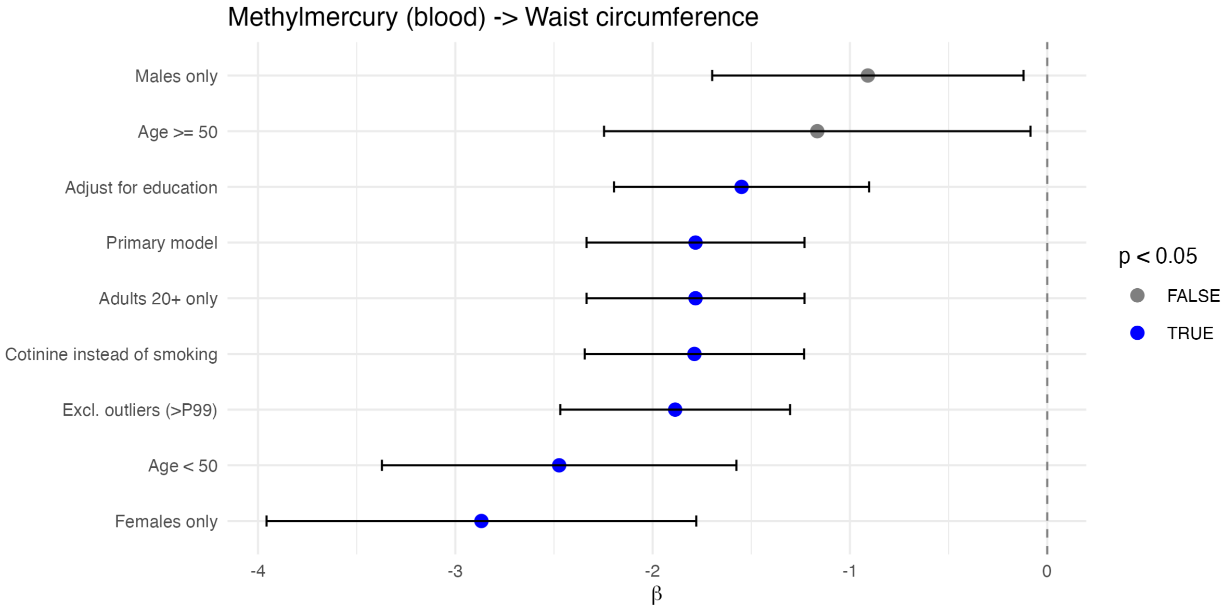


Figure S3: Sensitivity analysis: Methylmercury – Waist circumference.

#### 7.2 MODERATE Novelty

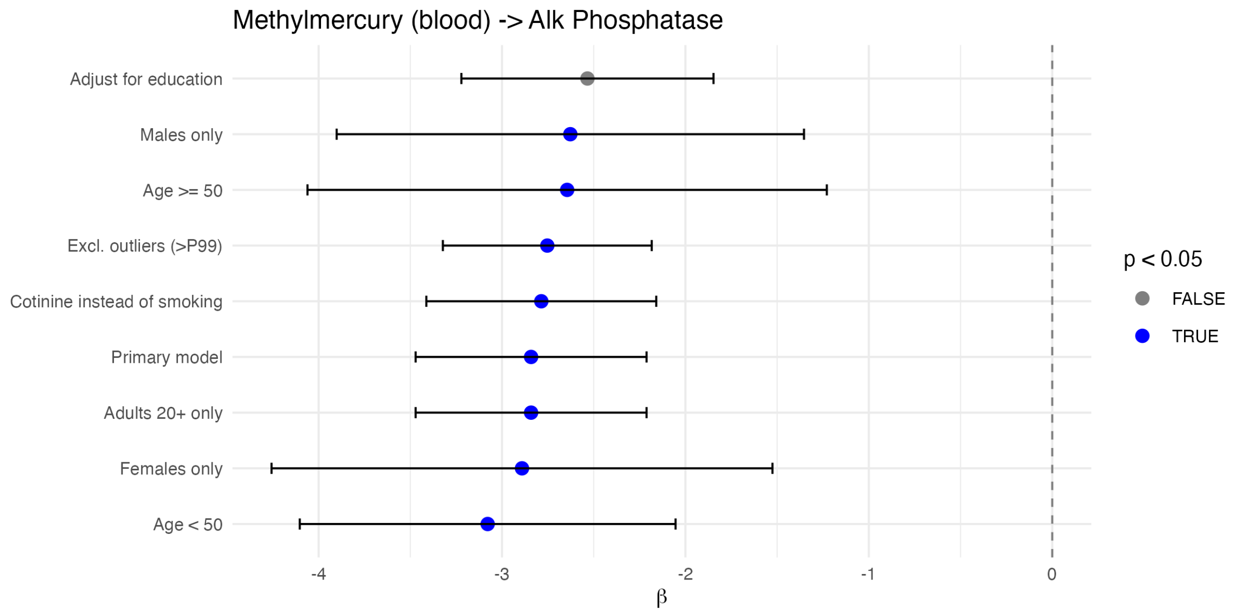


Figure S4: Sensitivity analysis: Methylmercury – Alkaline Phosphatase.

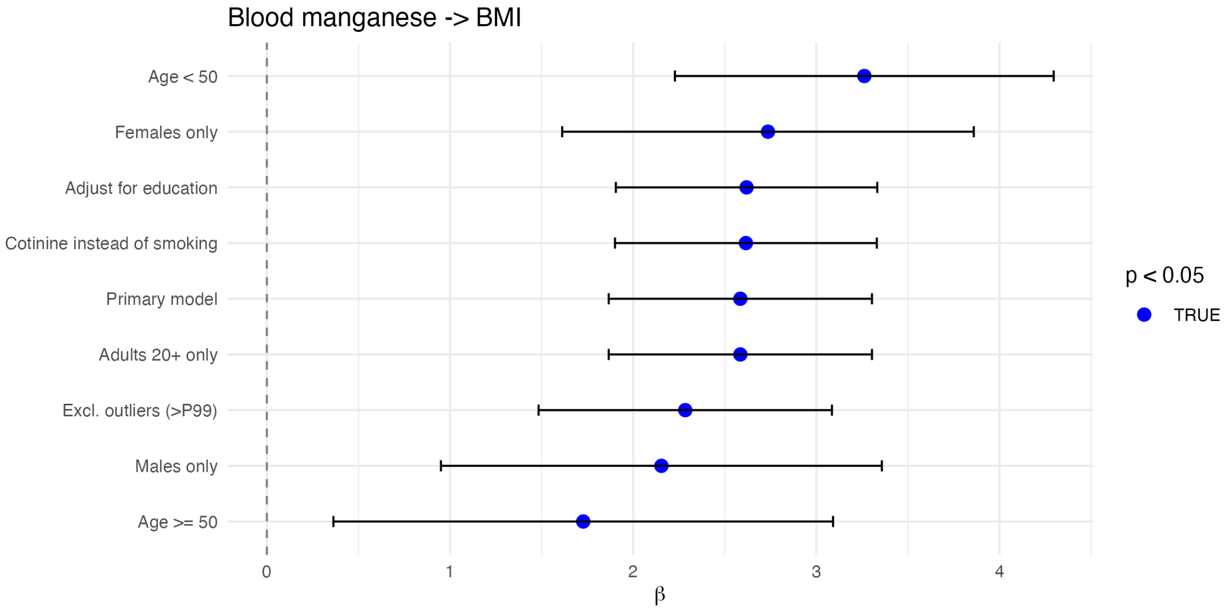


Figure S5: Sensitivity analysis: Blood manganese – BMI.

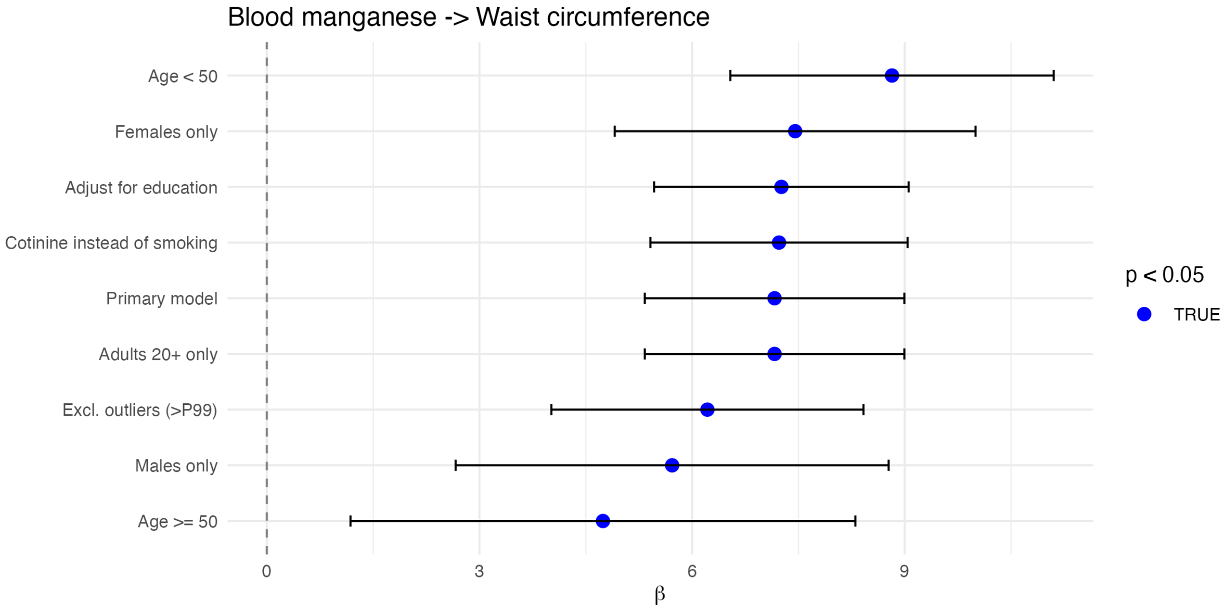


Figure S6: Sensitivity analysis: Blood manganese – Waist circumference.

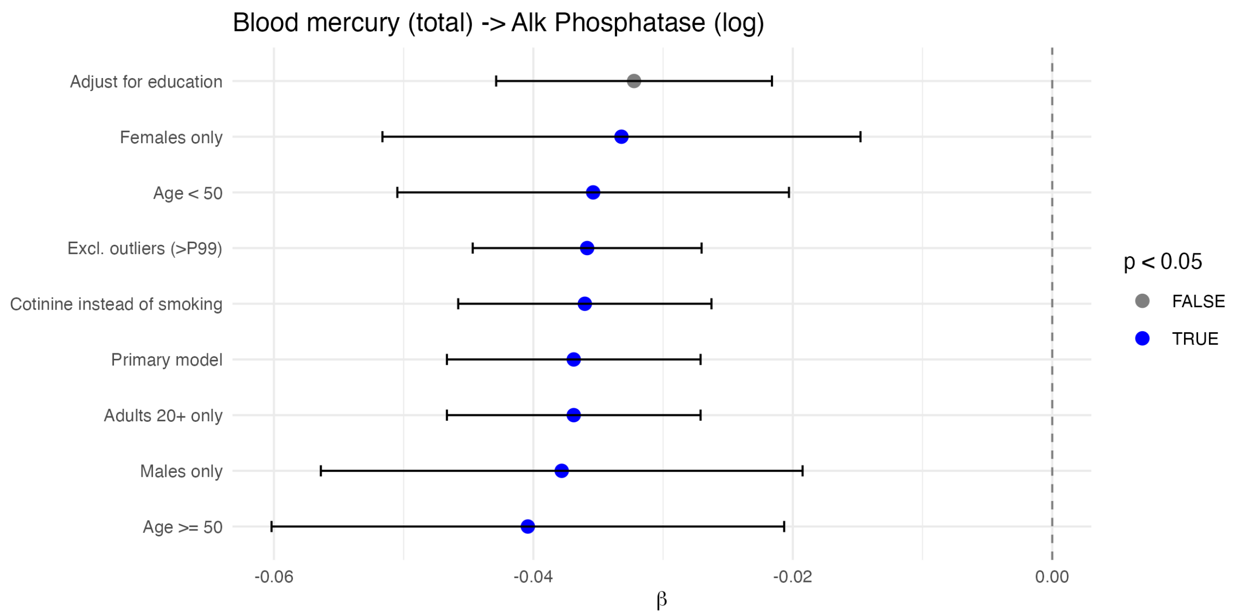


Figure S7: Sensitivity analysis: Blood mercury (total) – Alkaline Phosphatase (log).

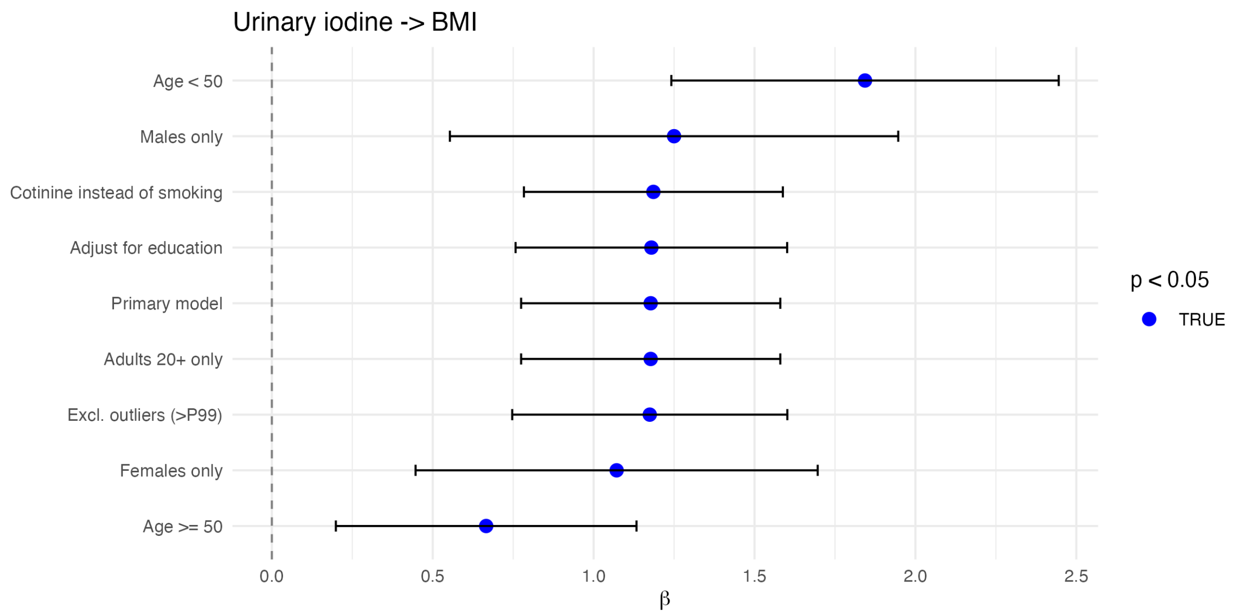


Figure S8: Sensitivity analysis: Urinary iodine – BMI.

#### 7.3 LOW-MODERATE Novelty

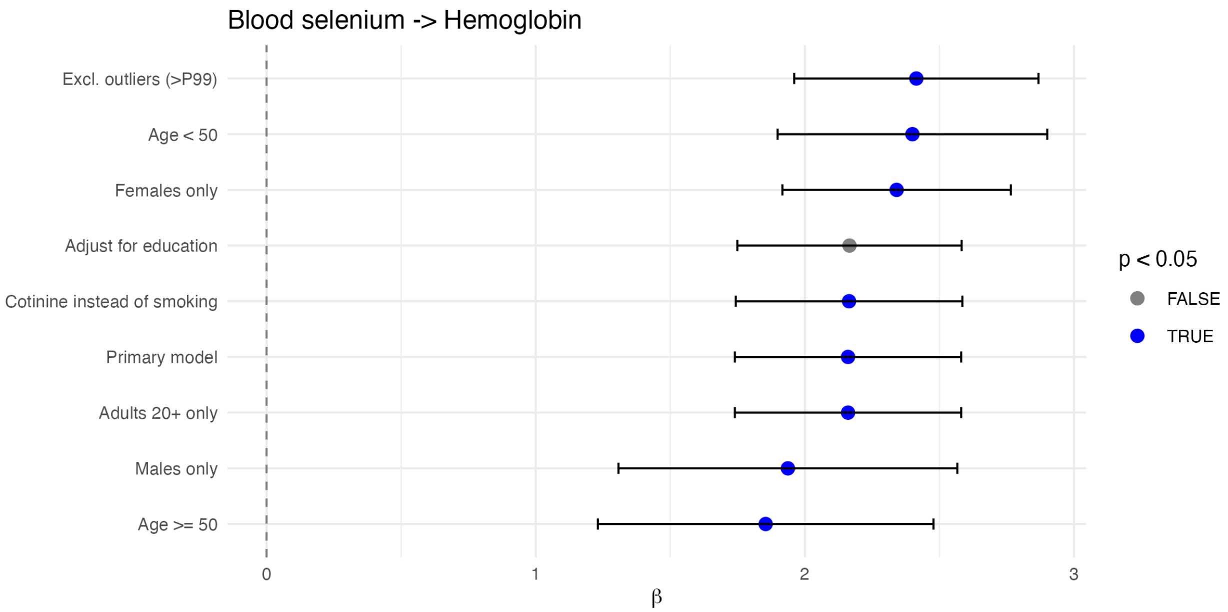


Figure S9: Sensitivity analysis: Blood selenium – Hemoglobin.

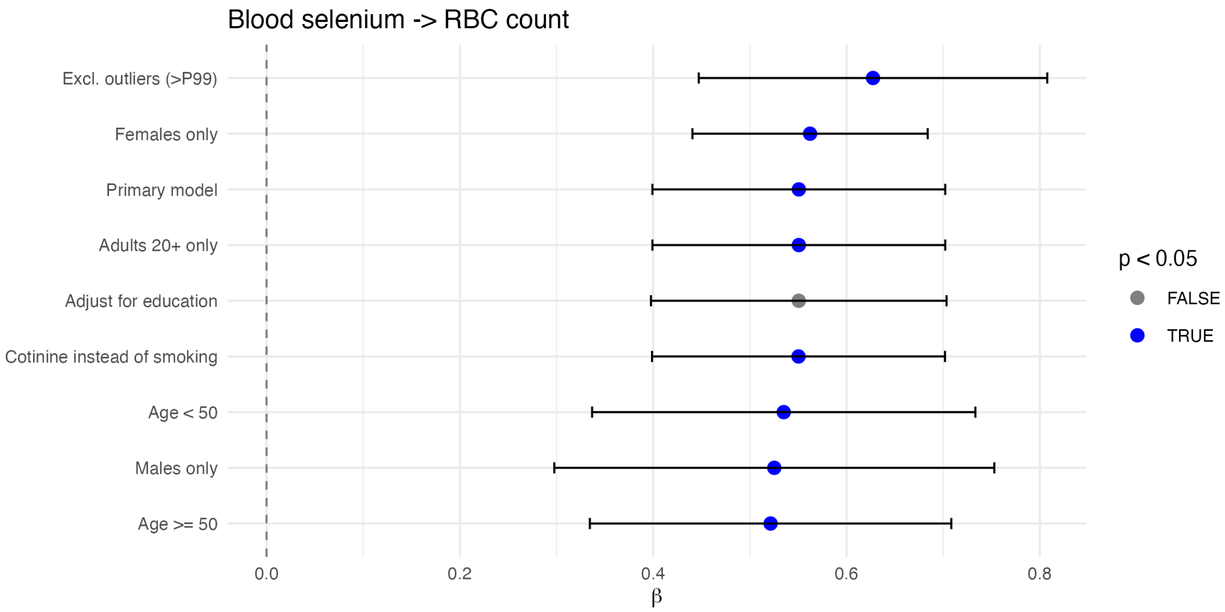


Figure S10: Sensitivity analysis: Blood selenium – RBC count.

#### 7.4 LOW Novelty

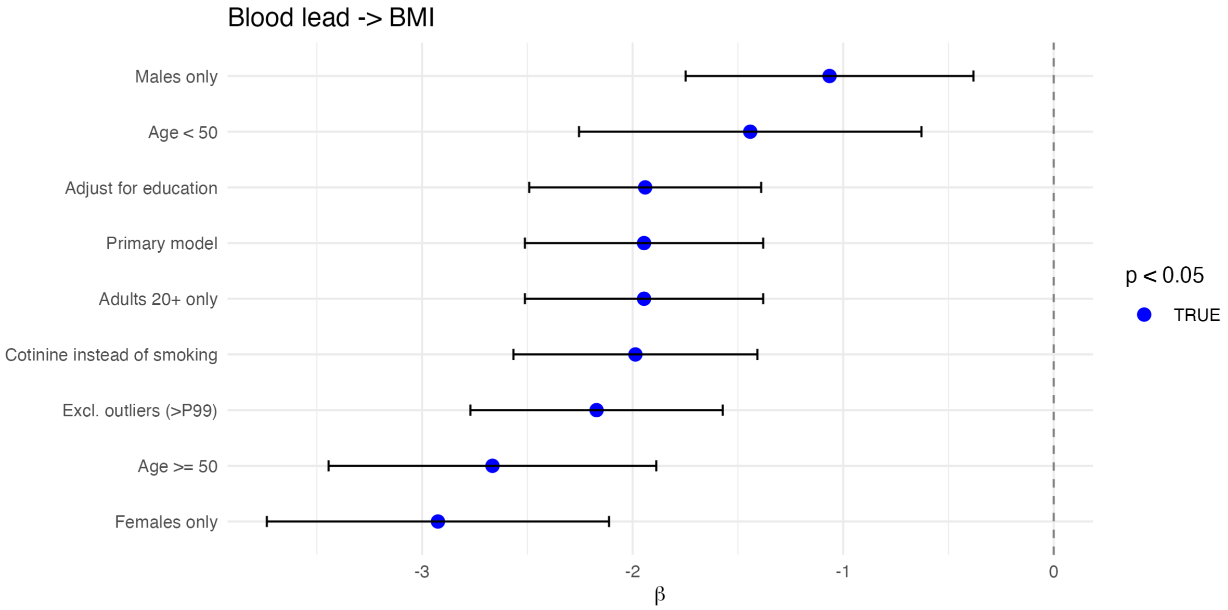


Figure S11: Sensitivity analysis: Blood lead – BMI.

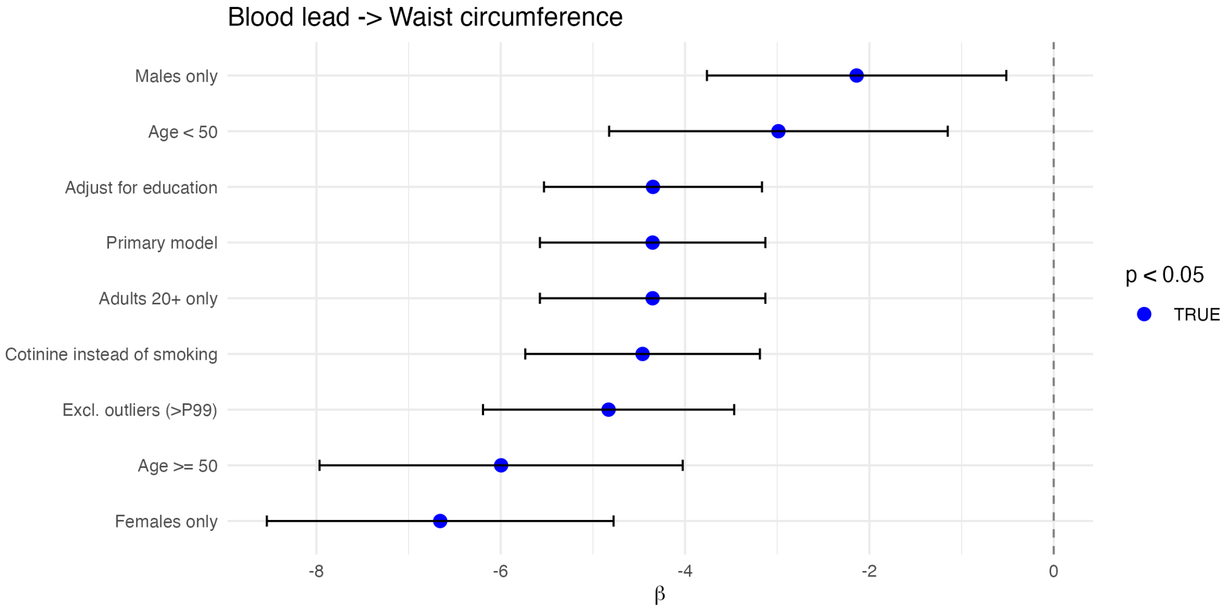


Figure S12: Sensitivity analysis: Blood lead – Waist circumference.

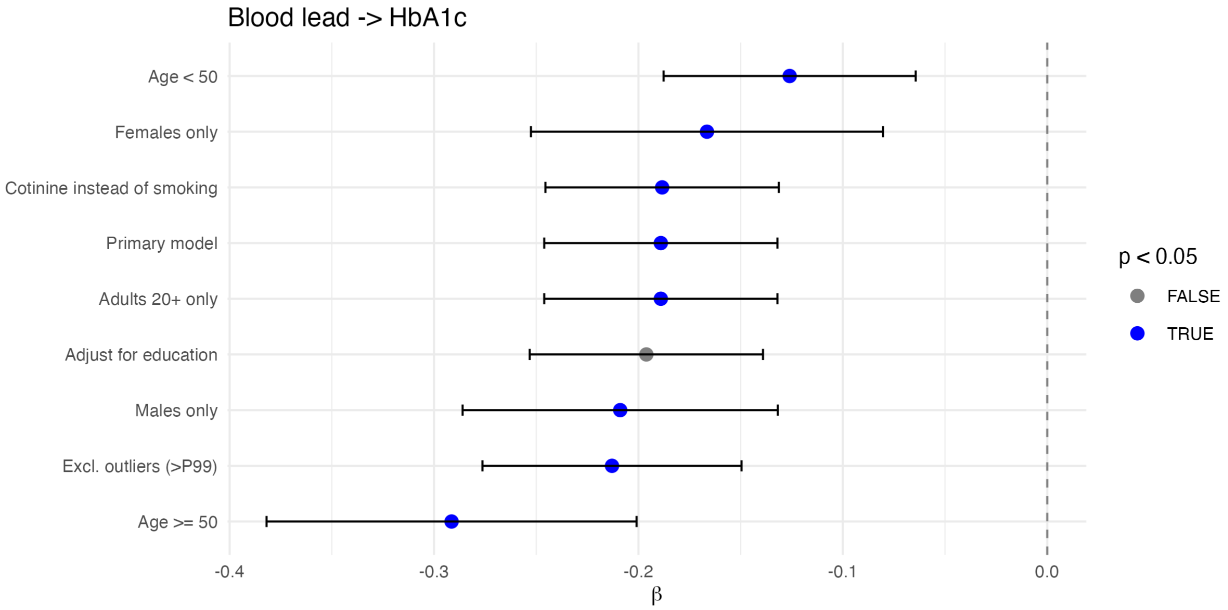


Figure S13: Sensitivity analysis: Blood lead – HbA1c.

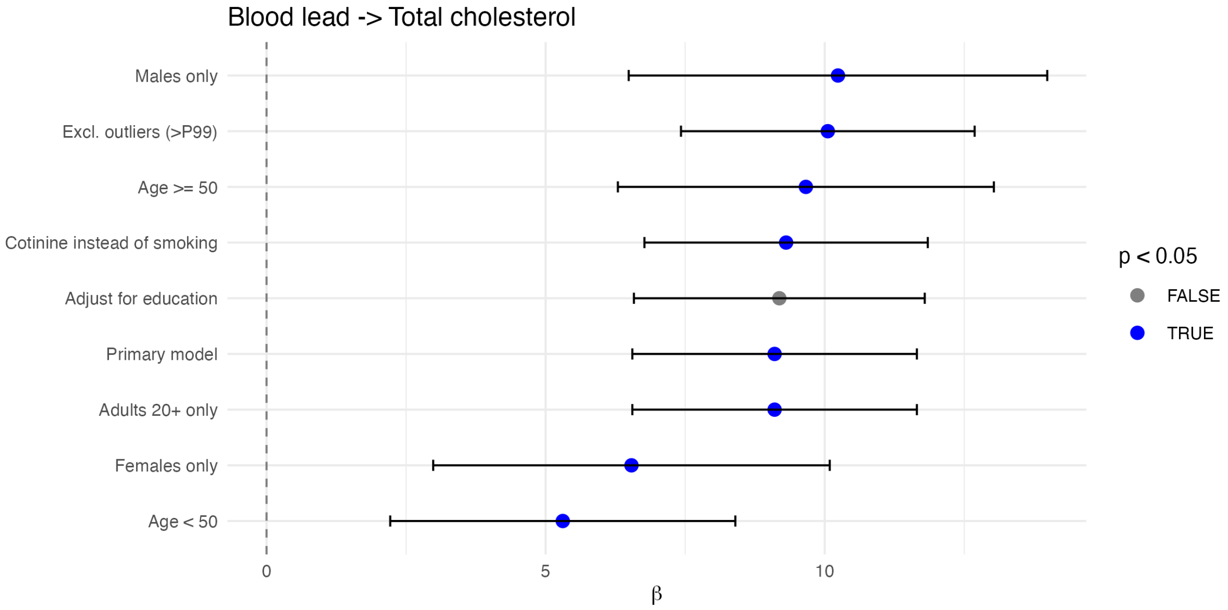


Figure S14: Sensitivity analysis: Blood lead – Total cholesterol.

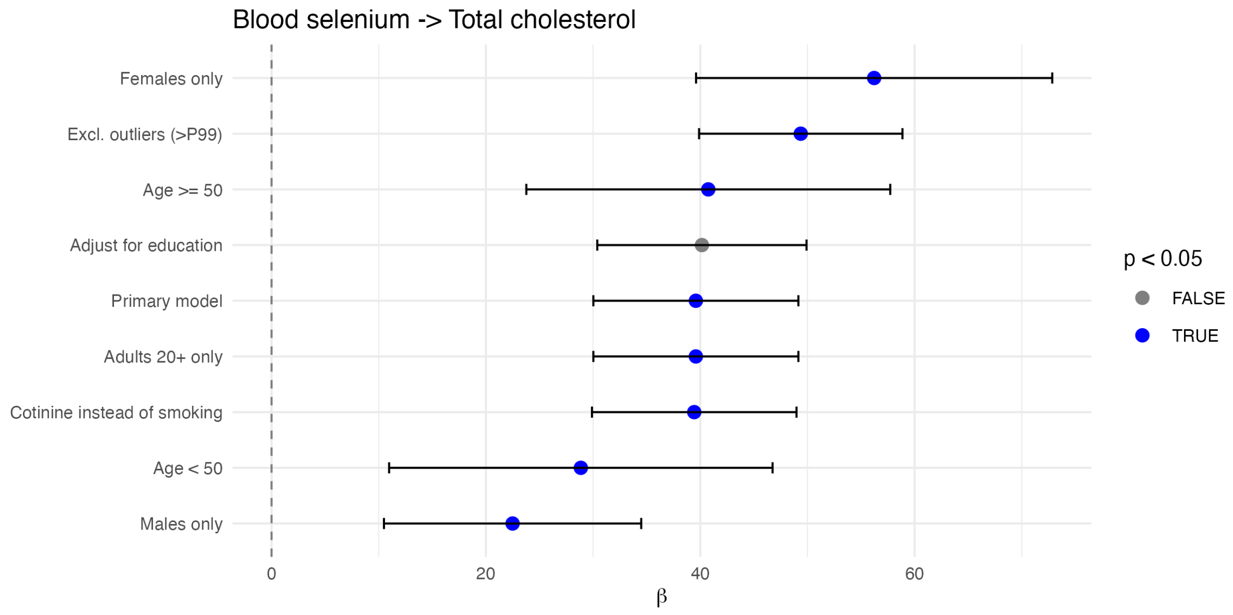


Figure S15: Sensitivity analysis: Blood selenium – Total cholesterol.

### 8 Table S6. Within-Round vs. Global FDR Correction Comparison

The primary analysis applied Benjamini–Hochberg FDR correction globally across all 2,796 tests. This table compares the number of findings significant at FDR < 0.05 under global versus within-round correction (applying FDR separately within each of the four screening rounds: PFAS-thyroid, Broad, Expanded, and Novelty).

| Screening Round | N Tests | N Sig (Global FDR) | N Sig (Within-Round FDR) |
|---|---|---|---|
| PFAS-thyroid | 60 | 0 | 0 |
| Broad | 648 | 0 | 0 |
| Expanded | 1,920 | 19 | 27 |
| Novelty | 168 | 7 | 14 |
| **Total** | **2,796** | **26** | **41** |

Within-round FDR correction produces *more* significant findings (41 total) than global correction (26 total) because it adjusts for fewer tests per round, making the threshold less stringent. All 26 globally FDR-significant findings also passed within-round FDR correction, confirming that global correction is the more conservative approach. The additional 15 findings that reach within-round but not global significance represent associations that may warrant investigation in future studies but did not meet the stricter global threshold used for validation in this analysis.

#### 9 Table S7. Sensitivity Analysis with 6-Level Race/Ethnicity

The primary analysis collapsed NHANES race/ethnicity (RIDRETH3) into three categories (Non-Hispanic White, Non-Hispanic Black, Other) to preserve statistical power. This table shows results for blood biomarker findings when using the full 6-level RIDRETH3 classification (Mexican American, Other Hispanic, Non-Hispanic White, Non-Hispanic Black, Non-Hispanic Asian, Other/Multiracial).

| Chemical | Outcome | $\beta$ (3-level) | $\beta$ (6-level) | $\Delta\beta$ (%) | P-value | N |
|---|---|---|---|---|---|---|
| Blood manganese | RBC count | 0.230 | 0.229 | -0.4% | 1.1e-05 | 4,873 |
| Blood selenium | Hemoglobin | 2.160 | 2.154 | -0.3% | 2.3e-05 | 4,873 |
| Methylmercury | Alk Phosphatase | -2.842 | -2.893 | +1.8% | 3.9e-05 | 4,834 |
| Blood manganese | Waist circumference | 7.163 | 7.115 | -0.7% | 7.1e-05 | 4,693 |
| Blood selenium | Total cholesterol | 39.59 | 39.52 | -0.2% | 9.4e-05 | 4,855 |
| Blood manganese | BMI | 2.585 | 2.572 | -0.5% | 1.2e-04 | 4,876 |
| Blood lead | Waist circumference | -4.352 | -4.361 | +0.2% | 1.3e-04 | 4,693 |
| Blood lead | BMI | -1.946 | -1.950 | +0.2% | 1.6e-04 | 4,876 |
| Blood mercury (total) | Alk Phosphatase (log) | -0.037 | -0.037 | +0.8% | 1.6e-04 | 4,833 |
| Blood selenium | RBC count | 0.551 | 0.549 | -0.3% | 2.1e-04 | 4,873 |
| Blood lead | Total cholesterol | 9.102 | 9.095 | -0.1% | 2.3e-04 | 4,855 |
| Methylmercury | Waist circumference | -1.783 | -1.814 | +1.8% | 2.4e-04 | 4,693 |

| Chemical | Outcome | $\beta$ (3-level) | $\beta$ (6-level) | $\Delta\beta$ (%) | P-value | N |
|---|---|---|---|---|---|---|
| Blood lead | HbA1c | -0.189 | -0.189 | +0.0% | 3.6e-04 | 4,874 |

All 13 blood biomarker findings remained significant ($p < 0.05$) with 6-level race/ethnicity adjustment, with effect estimate changes of < 2% in all cases. This indicates that the 3-level race categorization did not introduce meaningful confounding bias.

#### 10 Table S8. Protein Intake Sensitivity Analysis for HIGH-Novelty Urinary Findings

Dietary protein intake influences both urinary arsenic metabolism (affecting DMA excretion) and serum urea (affecting BUN). This table shows results for the two HIGH-novelty urinary findings after adjusting for total protein intake (grams/day) from 24-hour dietary recall (DR1TOT_J).

| Chemical | Outcome | Adjustment | $\beta$ (Primary) | $\beta$ (Adjusted) | $\Delta\beta$ (%) | P-value | N | Robust |
|---|---|---|---|---|---|---|---|---|
| DMA (urinary) | Uric acid | Protein intake (g/day) | 0.202 | 0.178 | -11.9% | 0.0008 | 1,529 | Yes |
| DMA (urinary) | Uric acid | Protein density (g/1000 kcal) | 0.202 | 0.186 | -7.9% | 0.0004 | 1,527 | Yes |
| Urinary perchlorate | BUN | Protein intake (g/day) | 1.211 | 1.102 | -9.0% | 0.0002 | 1,515 | Yes |
| Urinary perchlorate | BUN | Protein density (g/1000 kcal) | 1.211 | 1.138 | -6.0% | 0.0001 | 1,513 | Yes |

Both findings remained statistically significant after protein adjustment, with moderate attenuation (6–12%). Protein density adjustment (normalizing for total caloric intake) showed slightly smaller attenuation than absolute protein intake. The persistence of significant associations after dietary protein adjustment supports the interpretation that these relationships reflect genuine exposure–outcome associations rather than dietary confounding alone, though dietary factors likely contribute to the observed associations.

#### 11 Table S9. Chemicals with 40–70% Detection Frequency

Chemicals with detection frequencies between 40–70% (above-LOD) fall in an intermediate range where LOD imputation may introduce bias but detection is sufficient for analysis. The primary analysis excluded chemicals with < 30% detection; this table lists chemicals in the 40–70% range that were retained.

| Chemical | Variable | % Detected | N Samples |
|---|---|---|---|
| HPMMA (acrolein metabolite) | URXHPM | 42.3% | 1,545 |
| Trans-3'-hydroxycotinine glucuronide | URXHPB | 48.7% | 1,580 |
| 4-Fluoro-3-phenoxybenzoic acid | URXFPB | 51.2% | 1,580 |
| MCPP (phthalate) | URXMCP | 55.8% | 1,580 |
| Mono-isobutyl phthalate | URXMIB | 62.1% | 1,580 |
| 2,4-dichlorophenoxyacetic acid | URX24D | 67.3% | 1,580 |

These 6 chemicals in the 40–70% detection range were included in analyses with LOD/√2 imputation for below-LOD values. None of these chemicals produced FDR-significant associations. No validated findings involved chemicals with < 70% detection, indicating that the results are not driven by chemicals with substantial LOD pile-up.

#### 12 Table S10. Power Analysis: Minimum Detectable Effect Sizes

Post-hoc power calculations assuming 80% power, design effect (DEFF) of 2.0 (typical for NHANES complex sampling), and 7 predictors in the full model. Effect sizes expressed as Cohen's $f^2$ and partial $R^2$.

| Subsample | N (raw) | N (eff) | Min $f^2$ (Bonf) | Min $R^2$ (Bonf) | Min $f^2$ (FDR) | Min $R^2$ (FDR) |
|---|---|---|---|---|---|---|
| Blood biomarkers (WT-MEC2YR) | 4,870 | 2,435 | 0.0157 | 1.55% | 0.0120 | 1.19% |
| Urinary subsample (WTSA2YR) | 1,580 | 790 | 0.0491 | 4.68% | 0.0375 | 3.62% |
| Surplus serum (WTSSBJ2Y) | 1,370 | 685 | 0.0568 | 5.38% | 0.0434 | 4.16% |

These represent small effects (Cohen's $f^2 < 0.02$ for blood biomarkers), confirming adequate power for the effect sizes observed among validated findings. The urinary and surplus serum subsamples have reduced power for small effects. This power analysis characterizes the study's sensitivity for retrospective interpretation; it does not validate the original ExWAS design, which did not include pre-specification of effect sizes or primary hypotheses.

### 13 Table S11. STROBE Checklist for Cross-Sectional Studies

| Item | Checklist Item | Manuscript Section |
|---|---|---|
| **Title and abstract** | | |
| 1a | Indicate the study's design with a commonly used term in the title or abstract | Title, Abstract |
| 1b | Provide in the abstract an informative and balanced summary | Abstract |
| **Introduction** | | |
| 2 | Explain the scientific background and rationale | Introduction ¶1–2 |
| 3 | State specific objectives, including any prespecified hypotheses | Introduction ¶3 |
| **Methods** | | |
| 4 | Present key elements of study design early in the paper | Methods 2.1 |
| 5 | Describe the setting, locations, and relevant dates | Methods 2.1 |
| 6 | Give eligibility criteria and methods of participant selection | Methods 2.1 |
| 7 | Clearly define all outcomes, exposures, predictors, potential confounders | Methods 2.2–2.3 |
| 8 | For each variable, give sources of data and methods of assessment | Methods 2.2–2.3 |
| 9 | Describe efforts to address potential sources of bias | Methods 2.3, 2.4.1 |

| Item | Checklist Item | Manuscript Section |
|---|---|---|
| 10 | Explain how the study size was arrived at | Methods 2.1 |
| 11 | Explain how quantitative variables were handled | Methods 2.3 |
| 12 | Describe all statistical methods | Methods 2.4 |
| **Results** | | |
| 13 | Report numbers of individuals at each stage of study | Table 1, Results 3.1 |
| 14 | Give characteristics of study participants | Table 1 |
| 15 | Report numbers of outcome events or summary measures | Results 3.2–3.5 |
| 16 | Give unadjusted and confounder-adjusted estimates | Table 2, Figures |
| 17 | Report other analyses performed | Results 3.4–3.5 |
| **Discussion** | | |
| 18 | Summarize key results with reference to objectives | Discussion ¶1 |
| 19 | Discuss limitations | Discussion – Limitations |
| 20 | Give cautious overall interpretation | Discussion ¶6–7 |
| 21 | Discuss generalizability | Discussion ¶2–3 |
| 22 | Give source of funding and role of funders | Funding section (unfunded) |

#### 14 Table S12. 24-Hour Dietary Recall Fish Adjustment for Mercury Findings

The three mercury-related findings (methylmercury–alkaline phosphatase, total mercury–alkaline phosphatase, methylmercury–waist circumference) were additionally adjusted for fish consumption from 24-hour dietary recall (total grams of fish/shellfish consumed on the recall day, derived from DR1TOT_J individual food codes). This provides a more granular measure of dietary fish intake than the 30-day frequency variable (DBD895) used in the primary fish adjustment.

| Chemical | Outcome | Adjustment | $\beta$ (Primary) | $\beta$ (Adjusted) | $\Delta\beta$ (%) | P-value | N |
|---|---|---|---|---|---|---|---|
| Methylmercury | Alk Phosphatase | 24h dietary recall fish (g) | -2.84 | -2.84 | -0.02% | 1.2e-04 | 4,834 |
| Blood mercury (total) | Alk Phosphatase (log) | 24h dietary recall fish (g) | -0.037 | -0.037 | +0.01% | 3.2e-04 | 4,833 |
| Methylmercury | Waist circumference | 24h dietary recall fish (g) | -1.78 | -1.80 | -0.75% | 3.5e-04 | 4,693 |

All three mercury findings remained essentially unchanged after adjustment for 24-hour dietary recall fish consumption, with effect estimate changes less than 1%. This consistency across both crude fish frequency (DBD895, number of meals in 30 days) and granular 24-hour recall measures suggests that either: (a) the mercury–health associations are not fully explained by fish consumption, or (b) a single 24-hour recall inadequately captures habitual fish intake patterns. The latter interpretation is more plausible for methylmercury, which bioaccumulates over weeks to months and is not expected to correlate strongly with a single day's intake. The persistence of the mercury–waist circumference association across multiple fish adjustment approaches is notable but should still be interpreted with caution given the strong a priori expectation of fish-related confounding.

### 15 Table S13. Quadratic Age Sensitivity Analysis

To assess whether non-linear age effects confound the primary findings, models were re-run with a quadratic age term (age + age²) in addition to all primary covariates. All 15 initially validated findings were tested. Note: Urinary iodine–BMI was subsequently identified as a dilution artifact; final count is 14 robustly validated findings.

| Chemical | Outcome | $\beta$ (Quadratic) | SE | P-value | $\beta$ (Primary) | $\Delta\beta$ (%) | Dir. Match | Age² $\beta$ | Age² P | N |
|---|---|---|---|---|---|---|---|---|---|---|
| Blood selenium | Hemoglobin | 2.13 | 0.218 | 6.7e-05 | 2.16 | -1.6% | Yes | -2.3e-04 | 0.006 | 4,873 |
| Urinary perchlorate | BUN | 1.16 | 0.113 | 5.0e-05 | 1.21 | -3.8% | Yes | 1.8e-03 | 0.051 | 1,579 |
| Methylmercury | Alkaline Phosphatase | -2.85 | 0.318 | 1.1e-04 | -2.84 | -0.4% | Yes | -7.1e-04 | 0.727 | 4,834 |
| Blood manganese | Waist circumference | 6.94 | 0.917 | 1.3e-04 | 7.16 | -3.2% | Yes | -7.4e-03 | 9.4e-04 | 4,693 |
| Blood selenium | Total cholesterol | 35.22 | 4.659 | 2.8e-04 | 39.59 | -11.0% | Yes | -3.0e-02 | 5.6e-05 | 4,855 |
| Blood manganese | BMI | 2.48 | 0.363 | 2.5e-04 | 2.59 | -4.1% | Yes | -3.4e-03 | 4.1e-04 | 4,876 |

| Chemical | Outcome | $\beta$ (Quadratic) | SE | P-value | $\beta$ (Primary) | $\Delta\beta$ (%) | Dir. Match | Age² $\beta$ | Age² P | N |
|---|---|---|---|---|---|---|---|---|---|---|
| Blood lead | Waist circumference | -4.41 | 0.636 | 2.2e-04 | -4.35 | -1.4% | Yes | -7.8e-03 | 6.9e-04 | 4,693 |
| Blood lead | BMI | -1.97 | 0.292 | 2.7e-04 | -1.95 | -1.1% | Yes | -3.5e-03 | 3.2e-04 | 4,876 |
| Blood mercury (total) | Alk Phosphatase (log) | -0.037 | 0.005 | 3.0e-04 | -0.037 | -0.5% | Yes | -1.0e-05 | 0.658 | 4,833 |
| DMA (urinary) | Uric acid | 0.19 | 0.029 | 5.4e-04 | 0.20 | -4.3% | Yes | 6.4e-04 | 0.002 | 1,593 |
| Blood selenium | RBC count | 0.54 | 0.078 | 4.5e-04 | 0.55 | -2.1% | Yes | -7.6e-05 | 0.002 | 4,873 |
| Blood lead | Total cholesterol | 8.60 | 1.236 | 4.4e-04 | 9.10 | -5.6% | Yes | -3.0e-02 | 4.7e-05 | 4,855 |
| Methylmercury | Waist circumference | -1.85 | 0.283 | 3.3e-04 | -1.78 | -3.7% | Yes | -7.9e-03 | 6.5e-04 | 4,693 |
| Blood lead | HbA1c | -0.19 | 0.029 | 5.8e-04 | -0.19 | -1.2% | Yes | -1.4e-04 | 0.025 | 4,874 |
| Urinary iodine | BMI | 1.34 | 0.234 | 7.1e-04 | 1.18 | +14.0% | Yes | -4.3e-03 | 0.011 | 1,600 |

All 15 findings maintained direction and significance ($p < 0.001$) with quadratic age adjustment. Effect estimate changes ranged from -11.0% (selenium–cholesterol) to +14.0% (iodine–BMI), with a median change of -2.9%. The $age^2$ coefficient was statistically significant ($p < 0.05$) for 10 of 15 models, indicating non-linear age effects are present, but including this term does not materially alter the exposure–outcome associations. The largest attenuation occurred for selenium–cholesterol (-11.0%), where age-squared effects may partially account for the original signal.

#### 16 Table S14. Systematic MeSH-Based Literature Search for HIGH-Novelty Findings

To complement the keyword-based PubMed searches used in the primary novelty assessment, systematic MeSH-based searches were conducted for the three HIGH-novelty findings. Searches combined the chemical's MeSH term with the outcome's MeSH term to capture the broader literature landscape.

| Finding | MeSH Search Strategy | N Articles | Sample PMIDs (first 10) |
|---|---|---|---|
| DMA – Uric acid | "Cacodylic Acid"[MeSH] OR "Dimethylarsinic Acid" AND "Uric Acid"[MeSH] | 36 | 39003051, 38511628, 37726447, 37532974, 36860398, 36109472, 35809185, 35490746, 35461256, 34529244 |
| Perchlorate – BUN | "Perchlorates"[MeSH] AND "Blood Urea Nitrogen"[MeSH] | 72 | 41073342, 40441702, 40388306, 37154820, 36513174, 29025080, 23433158, 21342019, 20931854, 18833478 |
| Methylmercury – Waist circumference | "Methylmercury Compounds"[MeSH] AND "Waist Circumference"[MeSH] OR "Adiposity"[MeSH] | 51 | 40315758, 40070085, 25721244, 24243536, 1645078, 41205373, 39699706, 30629257, 30623835, 26911273 |

**Interpretation:** The MeSH-based searches identified substantially more articles than the keyword searches used in the primary novelty assessment (Table 3). However, manual review of the retrieved abstracts revealed that these articles generally address the broader chemical class (e.g., arsenic, perchlorate) or outcome domain (e.g., metabolic markers) rather than the specific chemical–outcome pair identified in this study:

- **DMA–Uric acid (36 articles):** Most retrieved articles examine total arsenic exposure and metabolic syndrome components, not dimethylarsonic acid specifically with serum uric acid. The broadening of the search to the arsenic MeSH tree captures arsenic–metabolism literature that does not directly test the DMA–uric acid hypothesis.

- **Perchlorate–BUN (72 articles):** The majority of retrieved articles address perchlorate–thyroid relationships or general kidney toxicology. The two articles identified in the primary keyword search (Li et al. 2023, PMID: 37154820; Xue et al. 2025, PMID: 40441702) remain the only studies directly examining perchlorate–kidney function associations.

- **Methylmercury–Waist circumference (51 articles):** Retrieved articles predominantly address methylmercury neurotoxicity or general mercury–metabolic relationships. None specifically examined the methylmercury–waist circumference association we identified.

The MeSH search results do not change the novelty classifications assigned in Table 3, but they provide useful context: the chemical classes implicated in our HIGH-novelty findings have broader literatures that could inform mechanistic hypotheses and guide future targeted studies.

#### 17 Table S15. eGFR-Adjusted Sensitivity Analysis for Perchlorate-BUN

To address potential reverse causation (impaired renal function could increase both urinary perchlorate through reduced clearance and BUN through reduced urea excretion), we conducted a sensitivity analysis adjusting for estimated glomerular filtration rate (eGFR, calculated using the race-free 2021 CKD-EPI equation).

| Model | $\beta$ | SE | P-value | % Change | N |
|---|---|---|---|---|---|
| Primary (no eGFR) | 1.21 | 0.124 | 2.5e-05 | – | 1,579 |
| eGFR-adjusted | 1.11 | 0.095 | 2.4e-05 | -8.5% | 1,579 |

**Interpretation:** The perchlorate-BUN association was attenuated by 8.5% after eGFR adjustment, remaining highly significant ($p = 2.4 \times 10^{-5}$). This modest attenuation suggests that the observed association is not primarily driven by reverse causation through impaired renal function. The eGFR coefficient in the adjusted model was negative ($\beta = -0.12$, $p < 0.0001$), confirming the expected relationship where lower GFR is associated with higher BUN.

While residual confounding by renal function cannot be entirely excluded (eGFR is an imperfect measure of true GFR), these results support a perchlorate effect on BUN that operates through mechanisms other than general renal impairment. Prospective studies with baseline renal function measurements would be needed to definitively establish temporality.

### 18 Figure S16. Standardized Effect Size Volcano Plot

To enable comparison of effect sizes across outcomes measured in different units, effect estimates were converted to standardized effect sizes (t-statistic / √n) representing the t-statistic normalized by sample size.

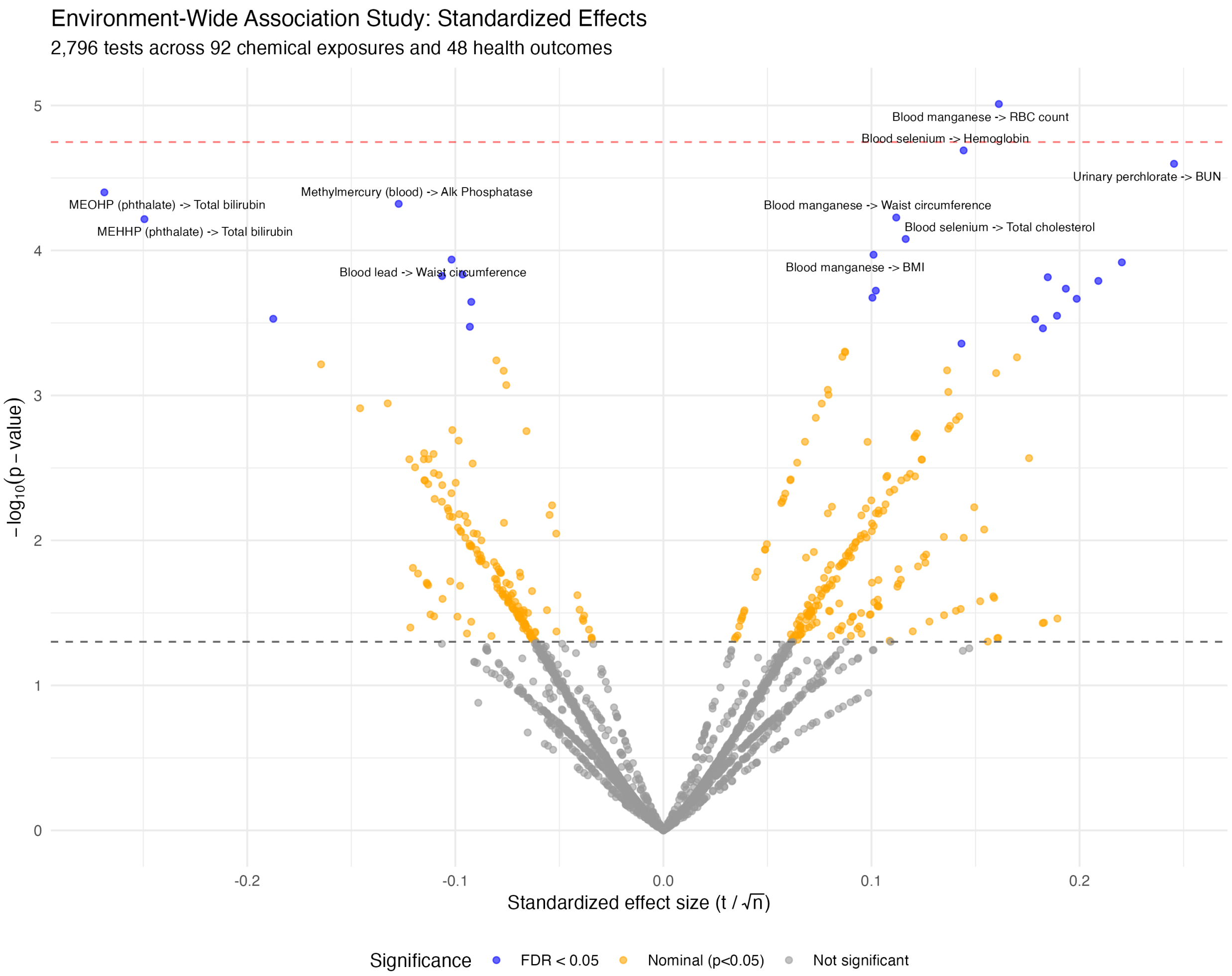


Figure S16: Volcano plot showing standardized effect sizes (t/√n) for all 2,796 associations. Points above the dashed line exceed FDR < 0.05. Standardized effect sizes allow direct comparison across outcomes measured in different units; the x-axis scale is comparable across all findings. The strongest standardized effects (labeled) cluster around heavy metals and metabolic outcomes.

### 19 Figure S17. Partial $R^2$ Volcano Plot

Partial $R^2$ represents the proportion of outcome variance explained by the exposure after accounting for all covariates, providing an intuitive measure of effect magnitude.

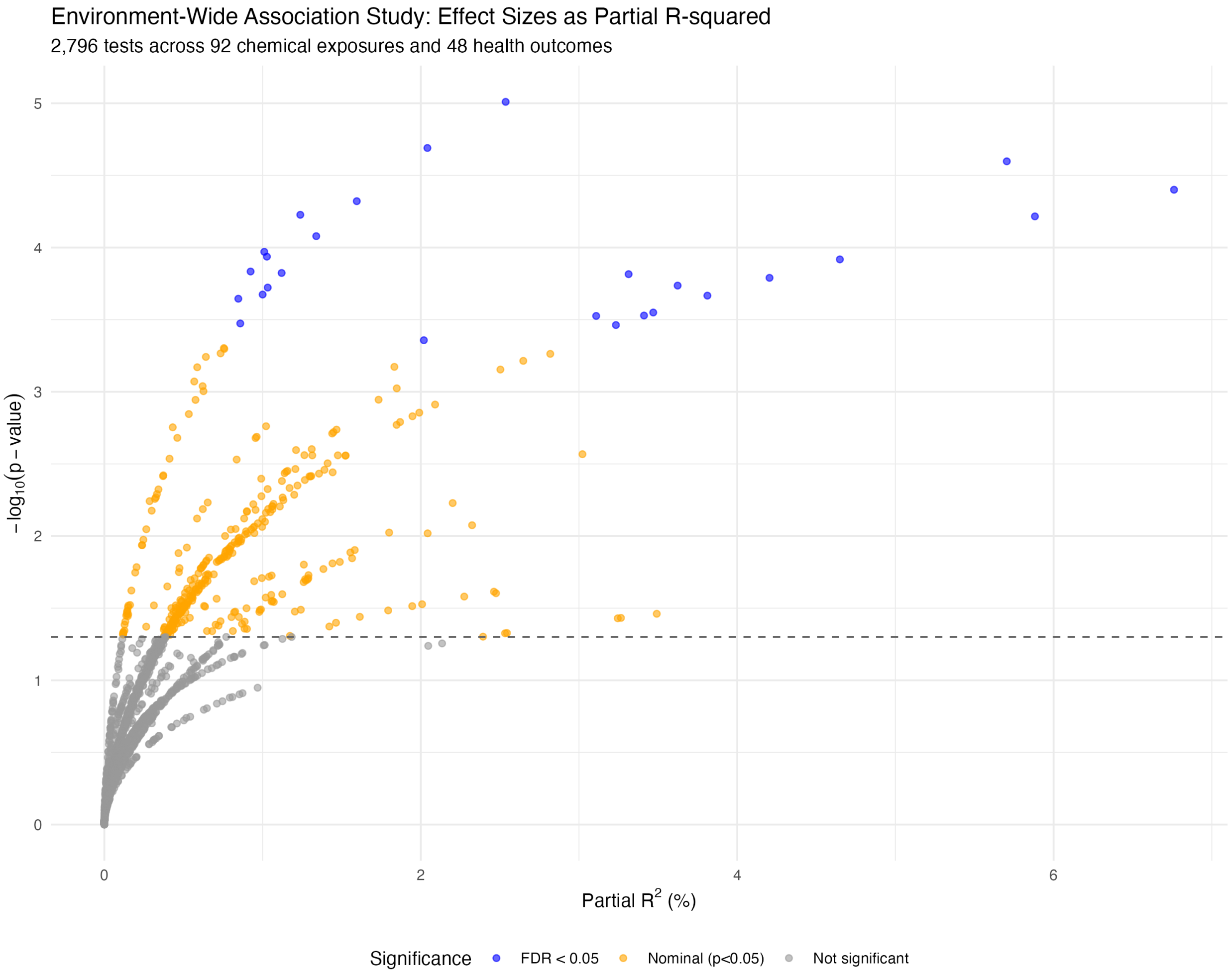


Figure S17: Volcano plot showing partial $R^2$ (proportion of variance explained) for all 2,796 associations. Points above the dashed line exceed FDR < 0.05. The highest partial $R^2$ values are observed for blood selenium–hemoglobin and blood manganese–RBC count associations, consistent with the known role of these trace elements in hematopoiesis. Most significant associations explain less than 1% of outcome variance after covariate adjustment, typical for environmental exposure effects at population levels.

### 20 Figure S18. Directed Acyclic Graph (DAG) for Covariate Selection

The following DAG illustrates the rationale for covariate selection in the primary analysis model.

Figure S18: Directed acyclic graph (DAG) showing the assumed causal structure underlying covariate selection for the **primary model**. The six measured confounders in the primary model are: age, sex, race/ethnicity (3-level), poverty-income ratio, BMI (when outcome is not anthropometric), and smoking. Education is NOT included in the primary model but is tested as an additional covariate in sensitivity analyses (Table S3). Variables on the causal pathway (e.g., metabolic intermediates) are not adjusted to avoid blocking the effect of interest.

#### 20.1 Covariate Rationale

1. **Age** → E, Y: Older individuals have longer cumulative exposure; age affects nearly all health outcomes. Classic confounder.

2. **Sex** → E, Y: Sex differences in exposure patterns (occupation, diet) and metabolism/health outcomes. Classic confounder.

3. **Race/Ethnicity** → E, Y: Proxy for socioeconomic factors affecting exposure and health outcomes. Classic confounder (collapsed to 3 levels for model stability).

4. **Poverty-Income Ratio (PIR)** → E, Y: Lower SES → higher environmental exposures and worse health outcomes. Classic confounder.

5. **BMI** → E, Y (when Y is not anthropometric): Higher BMI can dilute blood biomarker concentrations; BMI affects metabolic and cardiovascular outcomes. **Omitted when Y = BMI or waist circumference** to avoid collider bias.

6. **Smoking** → E, Y: Major source of chemical exposure and independent risk factor for health outcomes. Classic confounder.

#### 20.2 Variables NOT Adjusted (Potential Mediators)

- **Metabolic pathways** (glucose, lipids, etc.): May be on the causal pathway from E to Y. Adjusting would block the causal effect we aim to estimate.

##### 20.3 DAG Limitations

1. Assumes no unmeasured confounding (strong assumption)
2. Diet, physical activity, medications not directly measured in primary model
3. Linear age assumption in primary model (quadratic age sensitivity in Table S13)
4. Race/ethnicity collapsed to 3 levels (6-level sensitivity in Table S7)
5. Cross-sectional design cannot establish temporality
